# Performance and Challenges of Malaria Rapid Diagnostic Tests in Endemic Regions of Africa

**DOI:** 10.1101/2025.07.30.25331638

**Authors:** Fiyinfoluwa Demilade Ojeniyi, Adeola Oluwakemi Ayoola, Olajumoke Ibitoye, Oluyinka Oladele Opaleye, Olugbenga Adekunle Olowe, Leonard Ona Ehigie, Bolaji N. Thomas, Olusola Ojurongbe

**Affiliations:** Center for Emerging and Re-emerging Infectious Diseases, Ladoke Akintola University of Technology, Ogbomoso, Nigeria; Department of Biochemistry, Ladoke Akintola University of Technology, Ogbomoso, Nigeria; Department of Evolutionary, Anthropology, Duke University, Durham, North Carolina USA; Department of Computer and Information Sciences, Covenant University, Ota, Nigeria; Department of Medical Microbiology and Parasitology, Ladoke Akintola University of Technology, Ogbomoso, Nigeria; Department of Biomedical Sciences, College of Health Sciences and Technology, Rochester Institute of Technology, Rochester, NY 14623, USA

**Keywords:** Malaria, Rapid Diagnostic Tests (RDTs), Meta-analysis, Sensitivity, Specificity, Sub-Saharan Africa, Diagnostic Performance, HRP2 gene

## Abstract

Rapid diagnostic tests (RDTs) have revolutionized malaria diagnosis, playing a crucial role in improving timely treatment and supporting surveillance efforts, especially in resource-limited settings. However, the performance of RDTs can vary widely due to factors such as parasite genetic diversity, environmental conditions, and operational challenges. Understanding these variations is essential to ensuring accurate and reliable malaria diagnosis. This systematic review and meta-analysis critically evaluate the diagnostic performance of malaria RDTs across sub-Saharan Africa, identifying key gaps and proposing strategies for developing novel tests. By pooling data from 48 studies, the analysis quantifies the sensitivity and specificity of various RDT brands in different settings. The results reveal considerable variability, influenced by factors such as antigen persistence, cross-reactivity with other infections, and genetic polymorphism in the HRP2 gene, which can lead to false positives and negatives. The findings underscore the need for region-specific diagnostic strategies and the development of advanced diagnostic tools capable of detecting low-level parasitemia and differentiating between *Plasmodium* species. Emerging technologies and multi-platform approaches are recommended to enhance the accuracy and reliability of malaria diagnosis, ultimately contributing to more effective malaria control and elimination efforts in sub-Saharan Africa.

## INTRODUCTION

Accurate and rapid diagnosis is crucial for the timely and effective treatment of malaria, a significant public health challenge and potentially life-threatening disease in sub-Saharan Africa^1^, a disease induced by Plasmodium parasites transmitted to humans through the bite of an infected Anopheles mosquito. Rapid diagnosis is vital for effective monitoring and surveillance in public health programs^2^. The introduction of malaria rapid diagnostic tests (RDTs) in the mid-2000s has emerged as a promising solution, overcoming many challenges previously associated with achieving rapid, simple, and relatively affordable diagnostics for the disease^3^. RDTs are immunochromatographic assays that use lateral flow, where blood is applied directly to the test strip for antigen detection without requiring additional sample preparation or reagents. The test results are available within 15-30 minutes regardless of the absence of electricity or specific equipment. The extensive implementation of RDTs has significantly improved case management by ensuring that only confirmed malaria cases are treated, thereby reducing unnecessary use of antimalarial drugs and promoting confirmatory treatment based on accurate diagnosis. ^4^.

Despite the success of increasing diagnostic access with RDTs, several challenges and limitations necessitate ongoing monitoring and evaluation of these diagnostic tools^5^. Commercially available malaria RDTs typically detect either the Histidine-Rich Protein 2 antigen (HRP2), specific to *Plasmodium falciparum*, the Lactate Dehydrogenase (pLDH) antigen, common to all human malaria species, or the aldolase enzyme, which is also used as a target antigen in some RDTs.^6^. However, genetic polymorphisms and deletion in the *pfhrp2* and *pfhrp3* genes can affect the sensitivity and specificity of HRP2-based RDTs^7,8,10^. A study conducted in Nigeria showed that 17% of the analyzed isolates had deletions in the *pfhrp2* gene, and 6% had deletions in the *pfhrp3* gene. These deletions, particularly in the *pfhrp2* and *pfhrp3* genes, contribute to false negatives in RDTs, as their absence prevents the detection of histidine-rich proteins targeted by these tests. Another limiting factor in HRP-2 RDTs is the prozone effect, where samples with high parasite densities are occasionally undetected by RDTs^6,8^. Moreover, the sensitivity and specificity of RDTs vary significantly between commercial providers, compromising the diagnostic performance of these tests. This variation can affect the detection of different *Plasmodium* species, leading to inconsistent results across different test manufacturers and impacting the overall reliability of malaria diagnostics^9^. The ability of diagnostic tests to detect different malaria species and parasite life stages adds complexity, with some tests showing lower sensitivity for certain species or early life stages, leading to potential false negatives. This issue is significant in regions with diverse malaria species and varying parasite resistance levels^1^,.

The effectiveness of malaria diagnostics is significantly influenced by environmental conditions^11^ such as high temperatures and humidity, which can degrade RDT reagents, including components like the Lysis buffer, which may affect test performance. Additionally, high temperatures can denature the antibodies on the test strip, which are not classified as reagents but are critical for the proper functioning of the test. This degradation of both reagents and antibodies can reduce the accuracy and reliability of RDTs, posing a significant challenge to malaria control efforts in regions with variable environmental conditions.^12,13^. Misdiagnosis can occur in areas where HRP2-based RDTs that detect only *P. falciparum* are used, leading to false negatives for other malaria species^14^. Postinfection, persistent HRP2 antigens in the bloodstream can result in false positives, especially in high-transmission areas^6^. RDTs also lack sensitivity in detecting low-level parasitemia in asymptomatic or sub-microscopic infections, leading to missed diagnoses and delayed treatment^15^. The accuracy of malaria tests depends on the reference test, laboratory technician expertise, and established quality control procedures. Reference tests refer to confirmed diagnostic procedures, including microscopy or PCR, that check the accuracy of RDTs. Reference tests are standard for RDT performance evaluation during research, but field-based routine testing lacks these measures due to specialized equipment requirements and trained personnel needs. Using reference tests becomes feasible for reliability verification of RDT results, especially in high-burden areas or surveillance operations.^16^. Reports have indicated that malaria RDTs can cross-react with other infectious diseases and HRP2 can persist in the blood leading to false-positive results and unnecessary treatment^7^. This highlights the need to evaluate the specificity of RDTs in different epidemiological contexts and address false positive issues^17^. Considering the limitations of current RDTs and the ongoing challenges in malaria diagnostics and control, it is essential to identify diagnostic gaps and explore new frontiers for developing novel tests and improving existing ones^15^.

Addressing these challenges is crucial for reducing malaria’s impact, meeting ambitious global elimination goals, including reducing malaria mortality by 90% and eliminating the disease in 35 countries by 2030, and ensuring the long-term sustainability of malaria control efforts in sub-Saharan Africa. There’s an urgent need for optimized diagnostic tools that will be highly sensitive and specific while detecting low-level parasitemia and differentiating between Plasmodium species. These tools should not only be highly accurate but also user-friendly, especially for use at the point of care, where accessibility, simplicity, and rapid results are crucial for effective malaria management.^23^.

Molecular-based diagnostic tests, such as nucleic acid amplification tests (NAATs) and nextgeneration sequencing (NGS) technologies, offer higher sensitivity and specificity, with the potential to identify species and drug resistance genetic markers^16^. Notably, strategies for combining multiple diagnostic platforms or leveraging emerging technologies, such as microfluidics and biosensors, are being explored to enhance diagnostic accuracy and address the diverse challenges of current RDTs in different malaria-endemic regions^24,25^. Unfortunately, the high cost of setting up and maintaining these platforms in limited-resource countries and their limited use as point-of-care diagnoses are major disadvantages. Evaluating the performance of novel diagnostic tests is therefore crucial, considering factors such as local transmission patterns, parasite diversity, and operational feasibility in resource-limited settings^26^. Extensive field evaluations and implementation studies are still essential to ensure the successful deployment and integration of new diagnostic tools into existing malaria control programs^27^. Additionally, a comprehensive understanding of the current landscape of malaria RDT performance in sub-Saharan Africa is critical for identifying diagnostic gaps and informing strategies for the development of novel rapid tests^28^. This systematic literature review and meta-analysis aim to provide a rigorous synthesis and critical evaluation of the available evidence, offering insights into the strengths, limitations, and factors influencing the performance of existing RDTs in this region.

This review systematically analyzes published studies to (1) assess the diagnostic performance of malaria RDTs in sub-Saharan Africa, focusing on sensitivity, specificity, predictive values, target antigens, Plasmodium species, and transmission intensity; (2) identify diagnostic gaps and limitations; such as reduced sensitivity for detecting low-level parasitemia, false positives due to antigen persistence, and cross-reactivity; (3) examine geographical, epidemiological, and operational factors that influence RDT performance to enhance region-specific diagnostic strategies; and (4) provide insights and strategies for the development or enhancement of novel diagnostic tests, leveraging emerging technologies and multi-platform approaches tailored to the specific needs of malaria diagnosis in sub-Saharan Africa.

## RESULTS

### Search Results and Characteristics of the Included Studies

The comprehensive literature search identified a total of 678 records from various electronic databases. After removing 28 duplicate records, 650 unique records remained for screening. These records were assessed for eligibility, and 450 records were excluded for various reasons, including irrelevance to the study population, use of non-RDT index tests, and inappropriate reference methods. Detailed exclusion criteria are outlined in Figure 2 (PRISMA Flow Diagram).

The titles and abstracts of these records were carefully reviewed, excluding 450 records that did not meet the inclusion criteria. Consequently, 200 reports were identified for full-text retrieval and further assessment. All 200 reports were successfully retrieved and subjected to a thorough eligibility assessment. During this phase, 149 reports were excluded for various reasons: 50 reports were excluded because the study population was not relevant to the review question, 40 reports were excluded because the index test used was not an RDT, and 20 reports were excluded due to the use of inappropriate reference methods. Ultimately, 48 new studies met the predefined inclusion criteria and were included in the systematic review and meta-analysis. Of these, 41 studies focused on using RDTs to diagnose malaria, while 7 studies assessed the capacity of RDTs to monitor the effect of artemisinin-based combination therapy (ACT) treatment, with follow-up periods classified by days to evaluate the reduction in parasitemia. These studies provided a rich and diverse data set on the performance of malaria RDTs across different settings in sub-Saharan Africa. The distribution of these studies reflects a wide array of epidemiological contexts and malaria transmission intensities.

The geographical distribution map highlights the concentration of studies in West and Central Africa, particularly in Nigeria, Ghana, Cameroon, and Uganda (Figure 1A). Nigeria has the highest number of studies, with 11, Cameroon with 4, and Uganda with 3, as shown in Figure 1B. Ghana and Senegal each contributed 3 studies. This distribution underscores the focus on regions with high malaria transmission, providing a foundation for evaluating RDT performance in both high- and low-transmission settings. While data from high-burden countries offer valuable insights, incorporating data from diverse regions helps ensure a more comprehensive RDT reliability and accuracy assessment across different transmission environments.

**Figure 1:**
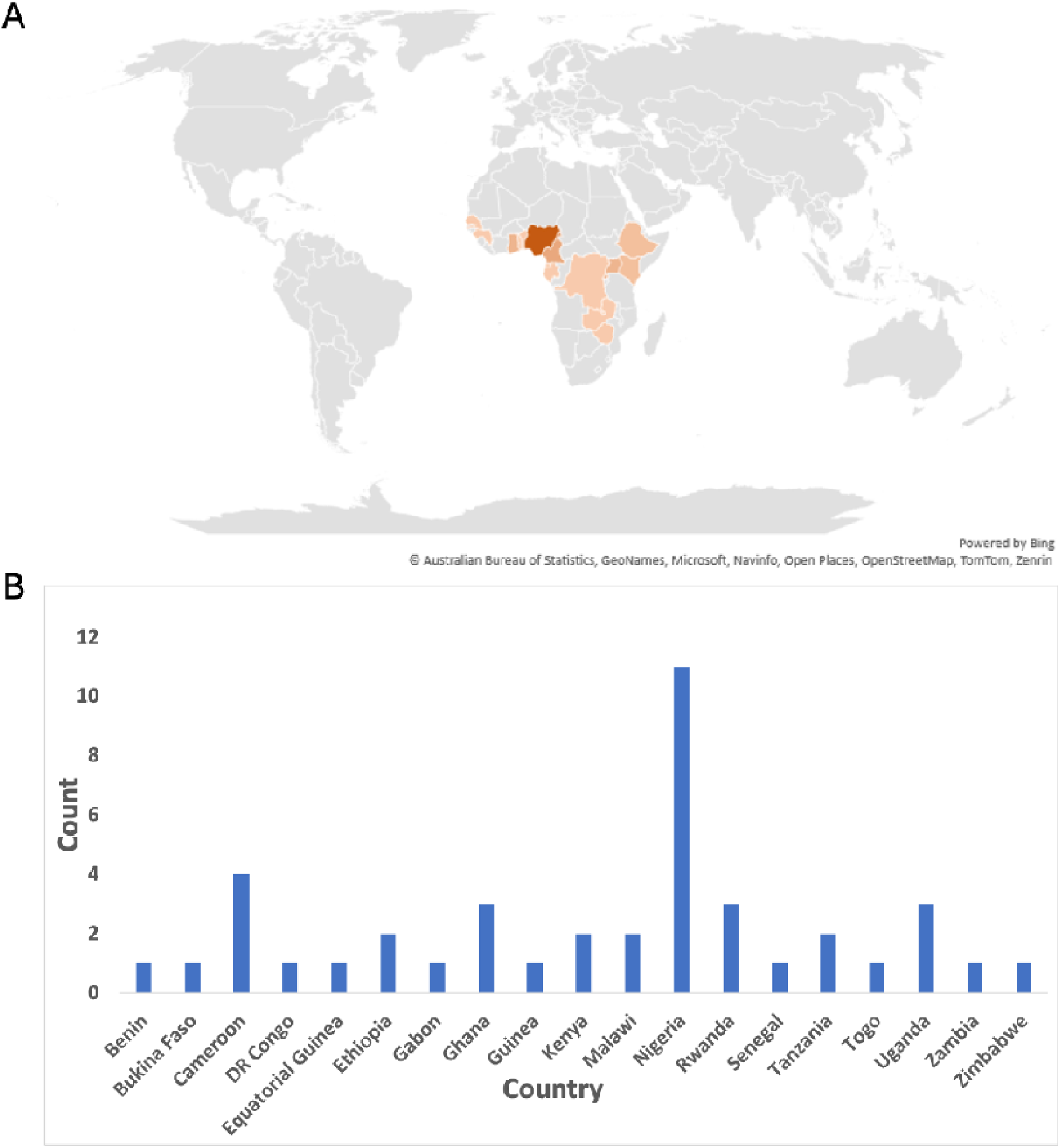
(A) Geographical Distribution Map of Included Studies. (B) Count of Studies by Country.

The PRISMA flow diagram (Figure 2) details the study selection process and the reasons for exclusion at each stage. This visual representation provides a clear overview of the identification, screening, and inclusion processes, ensuring transparency and reproducibility in the systematic review process.

**Figure 2:**
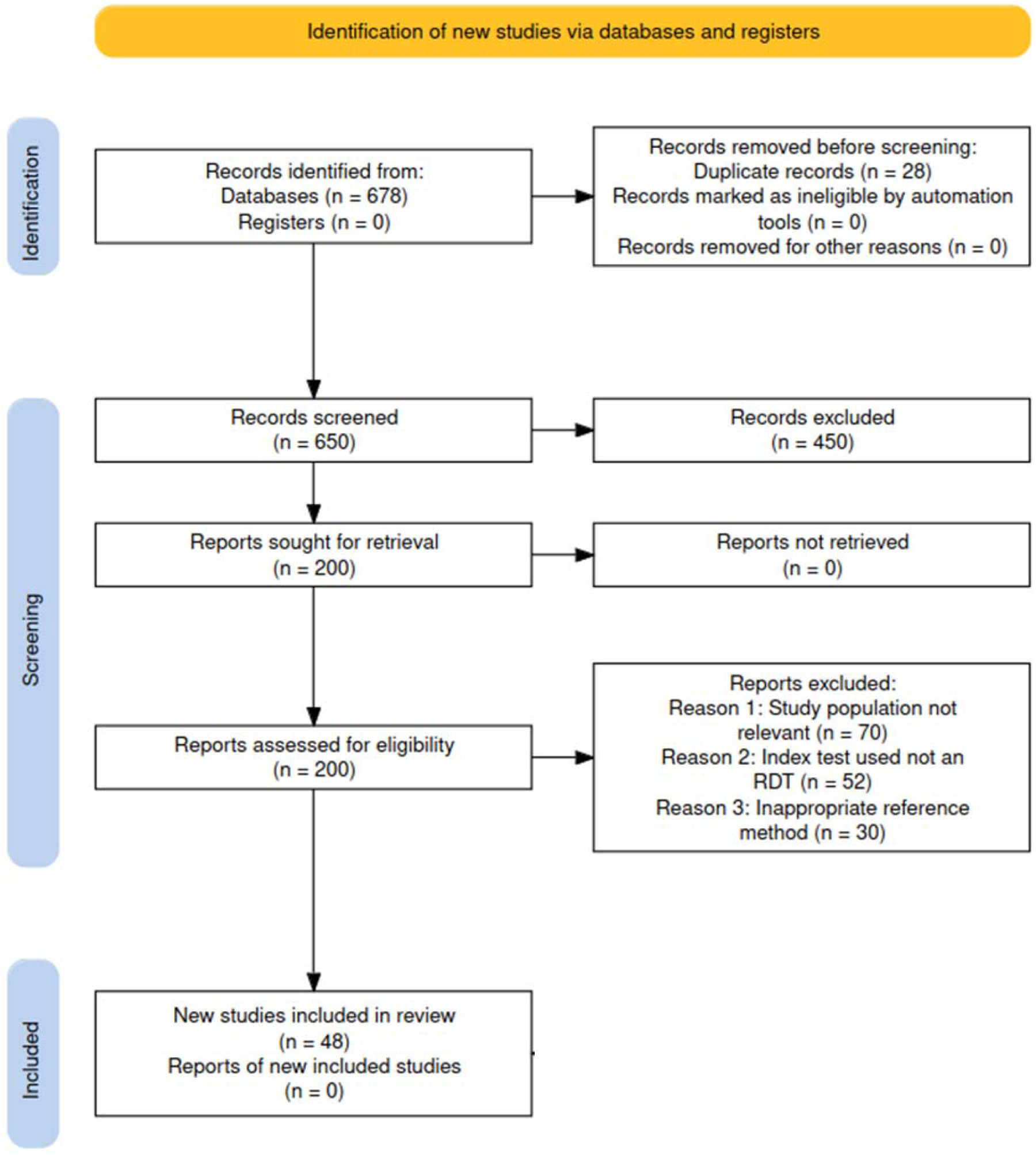
PRISMA Flow Diagram for Study Selection Process.

The systematic review and meta-analysis incorporated 48 studies conducted across various countries in sub-Saharan Africa. The studies included a mix of cross-sectional, diagnostic accuracy, and evaluation studies, highlighting the diverse methodologies employed to assess malaria RDTs. Sample sizes in the included studies varied significantly, ranging from as few as 32 participants to over 9,000, demonstrating both small-scale and large-scale research efforts. The studies encompassed a broad demographic spectrum, including infants (5 studies), children (12 studies), pregnant women (8 studies), symptomatic patients (18 studies), and asymptomatic patients (10 studies). The sex distribution and population characteristics varied, with some studies focusing on symptomatic individuals while others included asymptomatic participants.

Various malaria RDT brands and types were evaluated across the studies, with the following distribution: Type I (HRP2-based RDTs) was investigated in 25 studies, Type II (HRP2 and aldolase RDTs) in 8 studies, Type III (combined HRP2 and pLDH RDTs) in 7 studies, and ype IV (Pfspecific LDH; pan-specific LDH) in 6 studies. Brands evaluated in the studies included SD Bioline, CareStart, Paracheck, First Response, ICT Diagnostics, Gazelle, Alere, Advantage Malcard, and others. These variations in RDT types and brands highlight the diversity of diagnostic tools available and their respective performance in different settings. The reference standards used to validate the diagnostic accuracy of the RDTs included Giemsa-stained blood films microscopy, polymerase chain reaction (PCR), and composite tests.

The studies were conducted in diverse transmission settings, including 28 studies in high-transmission areas, 8 studies in moderate-transmission areas, and 12 studies in low-transmission areas, offering insights into the effectiveness of RDTs under different transmission intensities, from high to low, as well as seasonal transmission settings. The study periods ranged from 2010 to 2024, with data collection occurring across various seasons. This temporal diversity allowed for the evaluation of RDT performance during peak malaria transmission periods and throughout the year. Performance metrics, including sensitivity and specificity, varied among the studies. Some studies reported sensitivities and specificities above 90%, while others highlighted lower performance, particularly in detecting non*-P. falciparum* or low-density infections. Positive predictive value (PPV) and negative predictive value (NPV) were also reported, providing a comprehensive understanding of the diagnostic accuracy of RDTs in different settings.

Several studies reported limitations, such as the persistence of HRP2 antigenemia leading to false positives, variability in sensitivity and specificity across different brands and settings, and the impact of low parasite density on RDT performance. Additional challenges included cross-reactivity with other infections, the necessity for improved training and handling of test kits, and the importance of robust quality assurance measures. Overall, the included studies provided a comprehensive and diverse dataset on the performance of malaria RDTs. This information is crucial for understanding the current landscape of malaria diagnosis in sub-Saharan Africa and informing strategies for developing and improving diagnostic tools to enhance malaria detection and management in the region.

### Methodological Quality of the Included Studies

The methodological quality of the included studies was evaluated across four domains: patient selection, index test, reference standard, and flow and timing (Figure 3). Each study assessed the risk of bias and applicability concerns within these domains. For the patient selection domain, 94% (45 out of 48) of the studies demonstrated a low risk of bias, indicating the use of appropriate methods for selecting participants. However, 6% (3 studies) had concerns due to non-random sampling or specific inclusion criteria that might limit the generalizability of the findings. Most studies (94%) avoided inappropriate exclusions, ensuring a representative sample of the target population. The index test domain showed a predominantly low risk of bias, with 85% (41 out of 48) of the studies reporting clear procedures for conducting and interpreting the RDTs without knowledge of the reference standard results. However, 15% (7 studies) raised concerns due to the lack of blinding or pre-specification of thresholds, which could introduce bias in interpreting the index test results. For the reference standard domain, 90% (43 out of 48) of the studies demonstrated a low risk of bias, indicating that the reference standards used were appropriate for correctly classifying the target condition. Some concerns were noted in 10% (5 studies) related to interpreting reference standard results without knowledge of the index test results, potentially introducing bias. Nonetheless, overall, the reference standards were conducted and interpreted rigorously.

**Figure 3:**
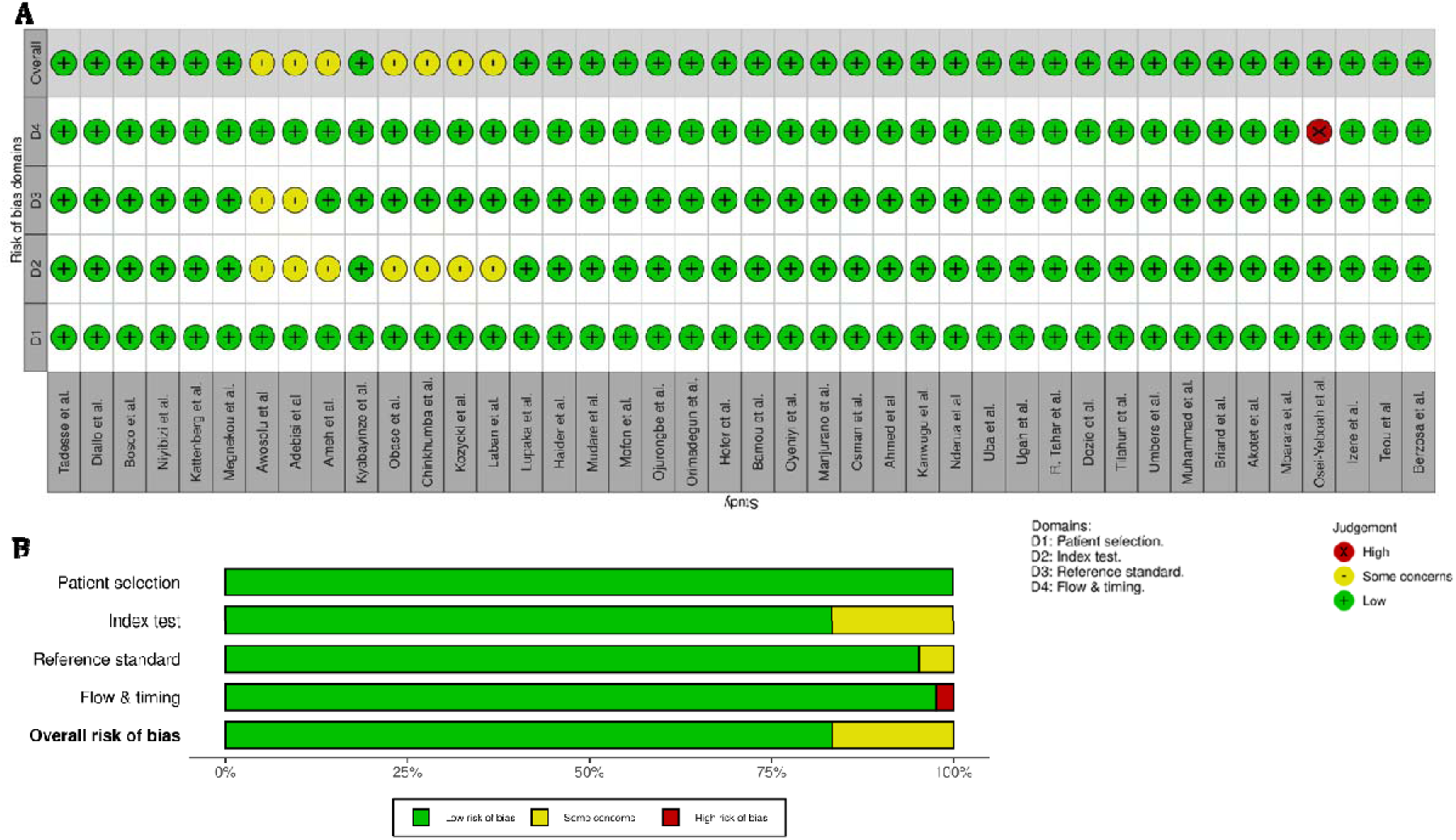
Risk of Bias Assessment Across Different Domains for Included Studies.

The flow and timing domain demonstrated a low risk of bias in 92% (44 out of 48) of the studies, which reported clear and appropriate intervals between the index test and the reference standard. Concerns in this domain were noted in 4 (8%) studies, primarily due to incomplete reporting of patient flow or timing issues that could affect the reliability of the findings. Nevertheless, most studies (92%) included all patients in the analysis. The overall risk of bias was low for most studies, with 40 (83%) out of 48 demonstrating low risk and 8 (17%) studies having some concerns, mainly in the index test and reference standard domains. Applicability concerns were generally low across all domains, indicating that the included studies were relevant to the review question and reflected realworld conditions in sub-Saharan Africa. Specifically, 47 (98%) out of 48 studies had low concerns regarding patient applicability, 43 (90%) out of 48 had low concerns regarding the index test applicability, and 44 (92%) out of 48 had low concerns regarding the reference standard applicability. The quality assessment using the QUADAS-2 tool indicates that the included studies were methodologically sound, with a few concerns primarily related to the interpretation of index tests and reference standard results. These findings provide confidence in the reliability of the diagnostic accuracy data reported in the studies and support the validity of the systematic review and meta-analysis conclusions.

### Overall Diagnostic Accuracy

The pooled sensitivity was found to be 0.830, with a 95% confidence interval (CI) ranging from 0.752 to 0.887, indicating a high probability of correctly identifying true positives. The specificity was determined to be 0.949, with a 95% CI between 0.913 and 0.971, demonstrating the tests strong ability to identify true negatives correctly. The false positive rate was relatively low at 0.051, with a 95% CI of 0.029 to 0.087 (Table 1). The analysis revealed a moderate negative correlation, as evidenced by the random effects correlation estimate -0.434. The diagnostic odds ratio (DOR) was 91.196, with a 95% CI ranging from 50.849 to 163.555, highlighting the high overall diagnostic accuracy of the tests. The likelihood ratios for positive and negative tests were 16.300 (95% CI: 9.676 to 27.459) and 0.179 (95% CI: 0.122 to 0.263), respectively, further supporting the efficacy of the tests (Table 1). Several critical aspects of the diagnostic performance were analyzed in detail. The value of theta (θ), estimated at -0.474, represents the overall mean log diagnostic odds ratio (log-DOR) across the studies, indicating the central tendency of diagnostic accuracy. A negative theta value suggests that, on average, the diagnostic performance is slightly below the expected level on the log-DOR scale. Lambda (λ), calculated at 4.414, reflects the precision of the random effects distribution, indicating the degree of variability in the log-DOR across the studies. A higher λ value suggests greater heterogeneity in diagnostic accuracy among the included studies. Beta (β), with a value of 0.174, accounts for the influence of covariates or moderators on the diagnostic performance. This coefficient adjusts the pooled estimates, ensuring that specific variables impacting diagnostic accuracy are considered. The variance components, sigma theta (σθ) and sigma alpha (σα) were estimated at 1.991 and 3.145, respectively, quantifying the variability in the log-DOR and the random effects. Sigma theta indicates the spread of the log-DOR across the studies, while sigma alpha captures the baseline variability in accuracy. The logit-transformed sensitivity was 1.588 (95% CI: 1.111 to 2.065), and the logit-transformed specificity was 2.925 (95% CI: 2.347 to 3.503). These transformations allow for linearized estimates, facilitating easier modeling and interpretation within the meta-analytic framework. Overall, the results underscore the robust and consistent diagnostic capabilities of the tests evaluated in this systematic review (**Table 1**).

**Table 1:** Diagnostic Performance Estimates for Malaria RDTs Across All Studies Included in the Systematic Review.

| Parameter | Estimate | 2.5% CI | 97.5% CI |
| --- | --- | --- | --- |
| Sensitivity | 0.83 | 0.752 | 0.887 |
| Specificity | 0.949 | 0.913 | 0.971 |
| False Positive Rate | 0.051 | 0.029 | 0.087 |
| Random Effects Correlation | -0.434 | - | - |
| $\theta$ | -0.474 | - | - |
| $\lambda$ | 4.414 | - | - |
| $\beta$ | 0.174 | - | - |
| $\sigma\theta$ | 1.991 | - | - |
| $\sigma\alpha$ | 3.145 | - | - |
| Diagnostic Odds Ratio | 91.196 | 50.849 | 163.555 |
| Likelihood Ratio (+ve) | 16.3 | 9.676 | 27.459 |
| Likelihood Ratio (-ve) | 0.179 | 0.122 | 0.263 |
| logit(sensitivity) | 1.588 | 1.111 | 2.065 |
| logit(specificity) | 2.925 | 2.347 | 3.503 |

Additionally, the Summary Receiver Operating Characteristic (SROC) curve (**Figure 4**) illustrates the diagnostic accuracy of malaria RDTs across multiple studies. The y-axis represents sensitivity, and the x-axis shows the false positive rate. Open circles denote individual study results, while the blue square indicates the pooled summary estimate. The dashed line outlines the 95% confidence region, and the dotted line represents the 95% predictive region, reflecting variability in test performance across different contexts.

**Figure 4:**
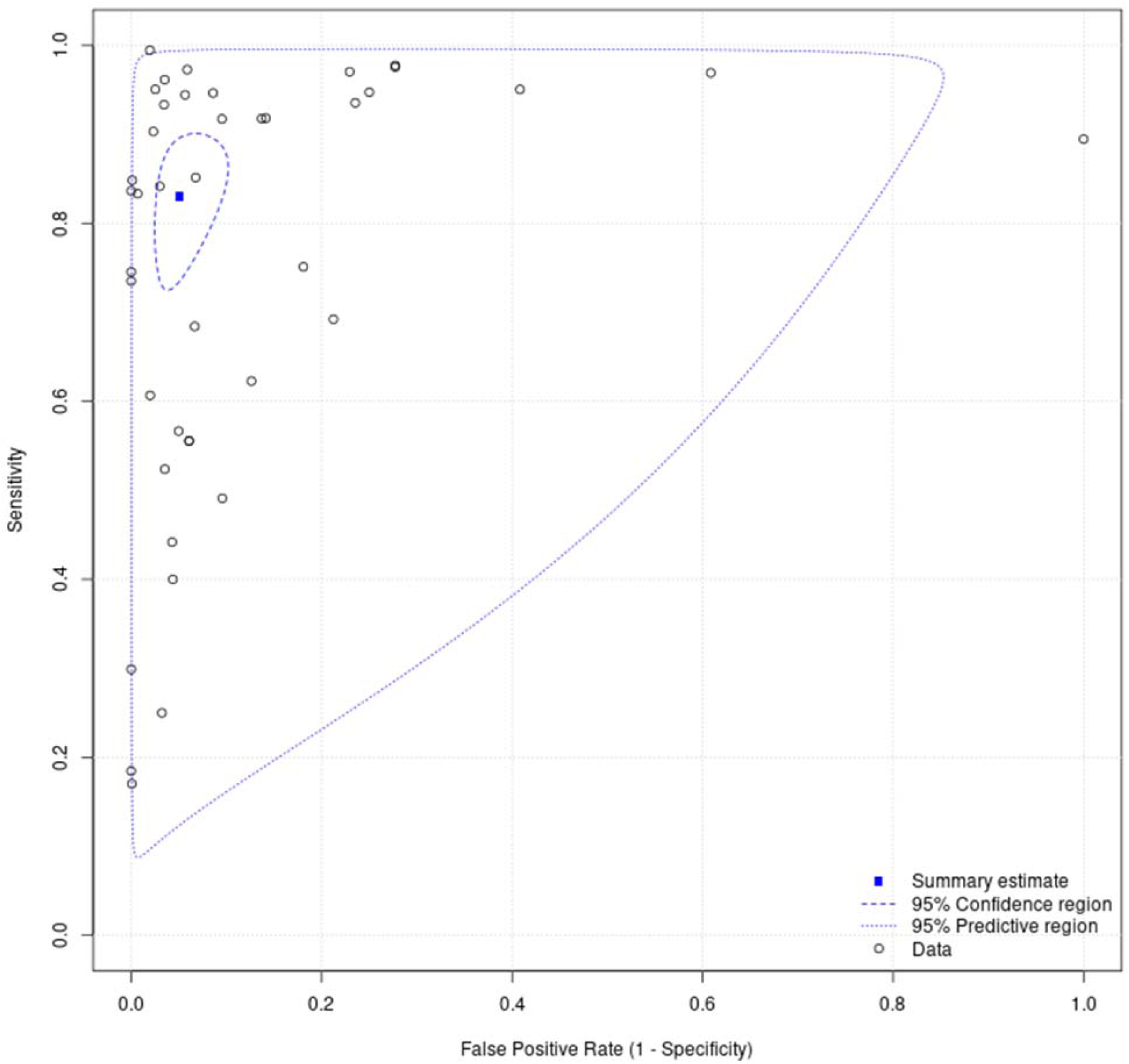
SROC Curve for Meta-Analysis of Malaria Rapid Diagnostic Tests (RDTs) *The SROC curve shows the diagnostic accuracy of malaria RDTs across multiple studies. The y-axis represents sensitivity, and the x-axis shows the false positive rate. Open circles denote individual study results, with the blue square indicating the pooled summary estimate. The dashed line outlines the 95% confidence region, while the dotted line represents the 95% predictive region, reflecting variability in test performance across different contexts*.

The analysis of malaria RDTs across multiple studies demonstrated high overall sensitivity and specificity, indicating their effectiveness in accurately identifying malaria cases. For example, a study in Ethiopia reported a sensitivity of 0.99 (95% CI: 0.97-1.00) and a specificity of 0.98 (95% CI: 0.93-1.00)^30^. Similarly, another study in Senegal recorded a sensitivity of 0.97 (95% CI: 0.93-0.99) and a specificity of 0.94 (95% CI: 0.86-0.98), further supporting the reliability of RDTs in different settings^31^. However, the performance of RDTs is not uniformly high across all studies. For instance, study conducted in Zambia reported a sensitivity of only 0.17 (95% CI: 0.08-0.31) despite a high specificity of 1.00 (95% CI: 1.00-1.00)^32^.

Regional analysis reveals significant differences in RDT performance attributed to local environmental and epidemiological factors. Studies from Ethiopia, Senegal, and Uganda generally report high sensitivity and specificity. For instance, two studies demonstrated high accuracy, suggesting that the RDTs used in these areas are well-suited to the local malaria strains and conditions^33,30^. Conversely, studies from Nigeria and Malawi indicate lower diagnostic accuracy potentially due to factors such as the local diversity of *Plasmodium* species, which might affect RDTs designed primarily to detect *Plasmodium falciparum*^34,35^. Environmental conditions like high humidity and temperature could degrade the test reagents, yielding unreliable results. These regional differences underscore the need for tailored diagnostic approaches considering local conditions and parasite diversity. The substantial variability among the included studies, as evidenced by the SROC analysis, can be attributed to differences in study design, sample sizes, population characteristics, and the specific brands and types of RDTs. Some studies exhibit high sensitivity and specificity, indicating consistent performance across different contexts^36,37^. In contrast, other studies show significantly lower sensitivity, highlighting potential issues with specific test implementations or local conditions impacting test accuracy^32^.

A random-effects model was used in the meta-analysis to account for variability between studies, ensuring that the sensitivity and specificity pooled estimates are robust and generalizable. Using a multivariate normal (MVN) distribution to model the logit-transformed sensitivity and specificity further strengthens the analysis. The parameter estimates (Table 2) for the bivariate normal distribution are as follows: logit(sensitivity) = 1.729, logit(specificity) = 3.081, σ²(logit(sensitivity)) = 0.090, σ²(logit(specificity)) = 0.090, and covariance = -0.032. These parameters provide a statistical foundation for understanding the relationship between sensitivity and specificity in the included studies.

**Table 2:** Summary of Heterogeneity Metrics.

| Metric | Estimate |
| --- | --- |
| Variance in Logit Sensitivity (Var logit(sen)) | 2.333 |
| Variance in Logit Specificity (Var logit(spe)) | 3.305 |
| Median Odds Ratio for Sensitivity (MOR sensitivity) | 4.292 |
| Median Odds Ratio for Specificity (MOR specificity) | 5.664 |
| Bivariate $I^2$ | 0.733 |
| Area of 95% Prediction Ellipse | 0.47 |

The forest plots for sensitivity and specificity provide a clear visual representation of the variability and overall performance of RDTs across different studies. The forest plot for sensitivity (Figure 5) illustrates that while many studies report high sensitivity, significant outliers exist with much lower sensitivity values. This contrasts sharply with the high sensitivity reported in some studies^30^.

**Figure 5:**
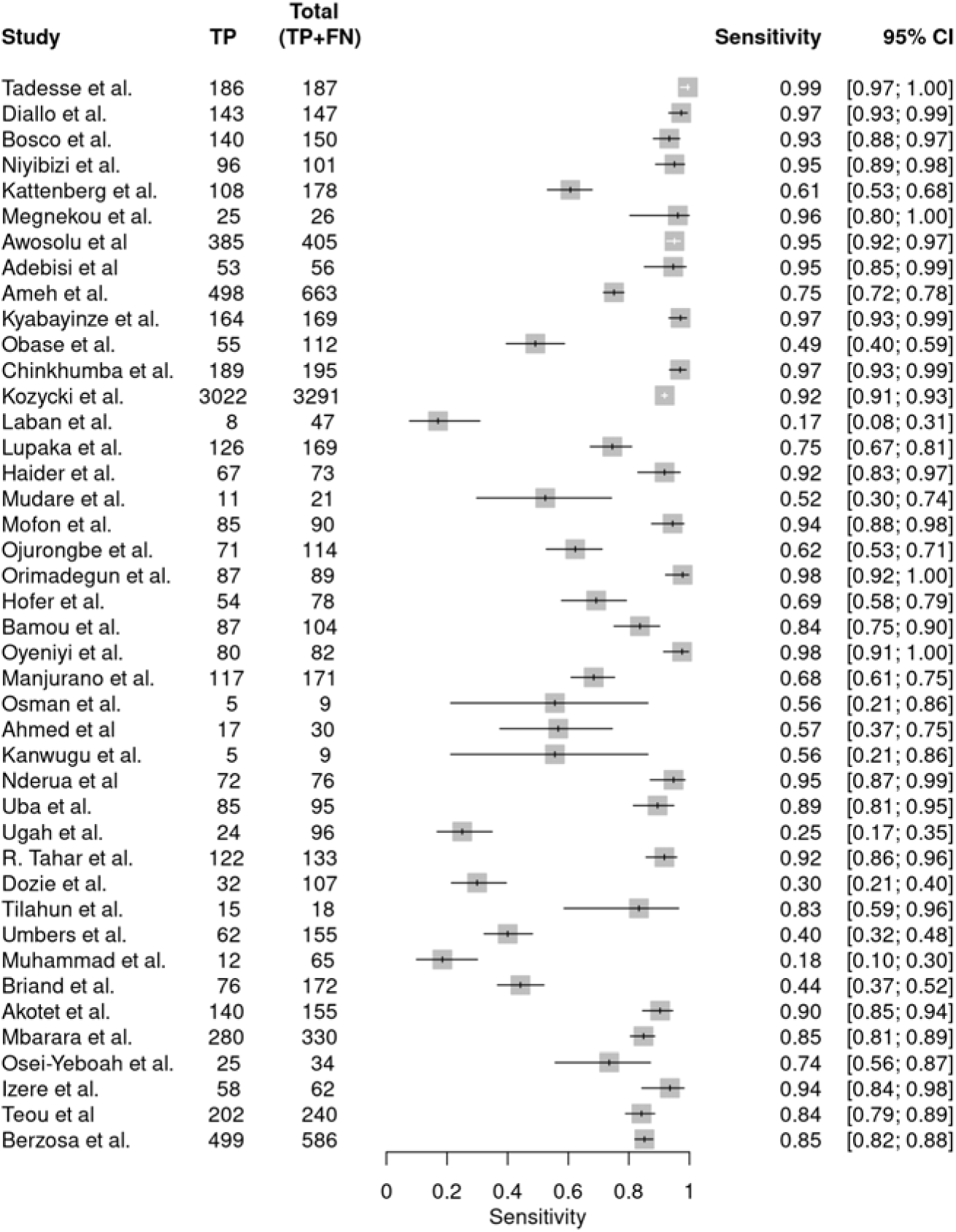
Forest plot of sensitivity.

Similarly, the forest plot for specificity (Figure 6) shows generally high specificity across studies, with some variability. Most studies report high specificity, indicating that RDTs effectively identify individuals who do not have malaria^31,36^. However, the lower specificity observed in other studies suggests the presence of false positives^34,38^. These visual aids are crucial for understanding the trade-offs between sensitivity and specificity in different settings. They highlight the importance of considering both measures when evaluating the diagnostic performance of RDTs and guiding future research and implementation strategies.

**Figure 6:**
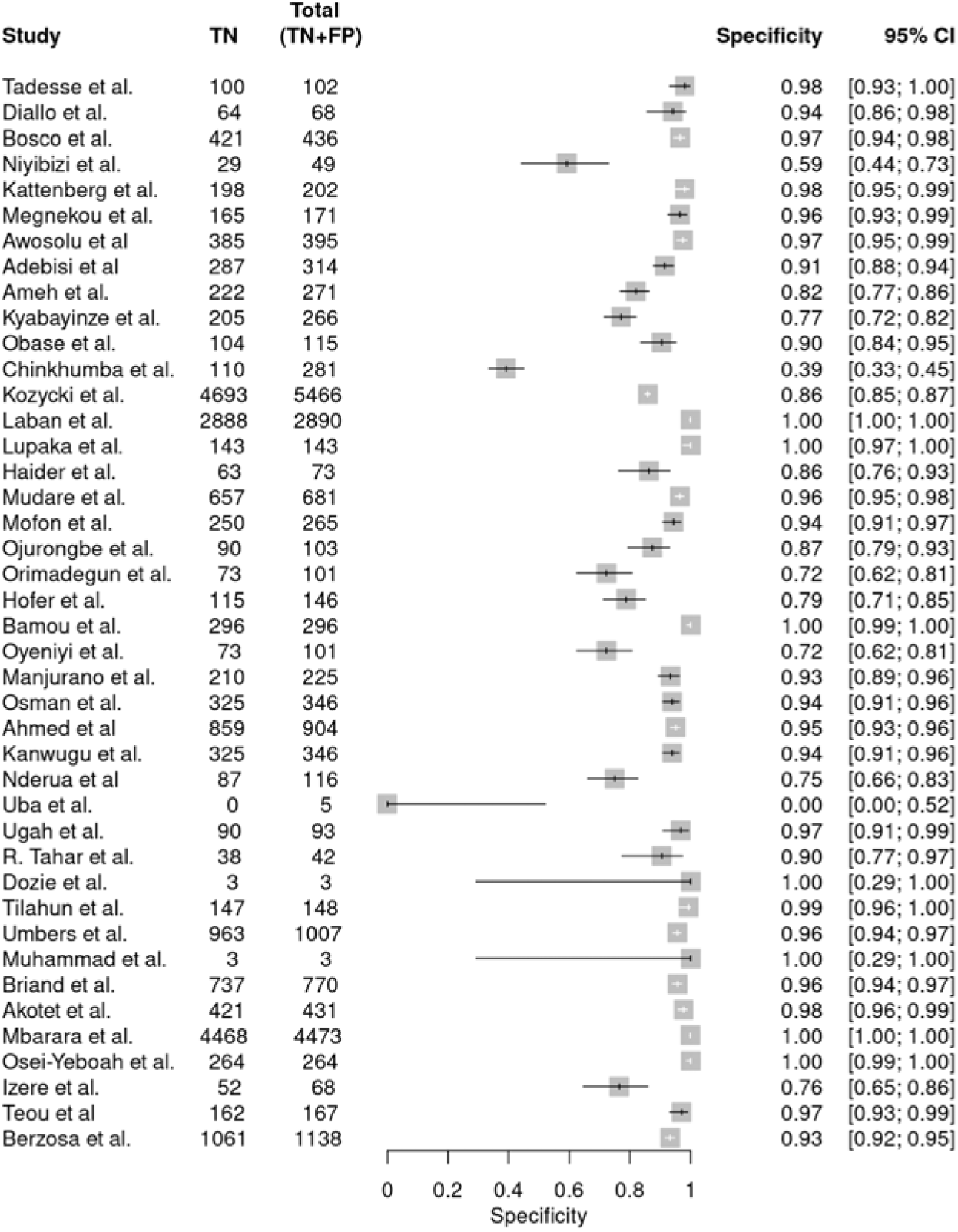
Forest plot of specificity.

### Patterns and Gaps in Diagnostic Accuracy Data

Analysis of the sensitivity and specificity data from various studies on malaria RDTs reveals several key patterns and gaps in diagnostic performance across different study designs and regions. While many studies demonstrate high sensitivity and specificity, indicating effective diagnostic performance^30^, variability exists due to study design, geographical region, and local conditions. Some studies report consistently high diagnostic accuracy^31^, while others show discrepancies, especially in regions with diverse *Plasmodium* species or challenging environmental conditions^33^. Despite the generally high diagnostic performance, variability in sensitivity exists among the studies. For instance, one study reported a notably low sensitivity of 0.17 (95% CI: 0.08-0.31), despite a high specificity of 1.00 (95% CI: 1.00-1.00), indicating potential issues with test performance under certain conditions or populations, possibly due to variations in parasite density, test administration, or environmental factors^32^. Another study reported moderate diagnostic accuracy with a sensitivity of 0.61 (95% CI: 0.53-0.68) and specificity of 0.98 (95% CI: 0.95-0.99), potentially influenced by local factors such as parasite strain differences or operational challenges^39^. Another study showed a sensitivity of 0.75 (95% CI: 0.72-0.78) and specificity of 0.82 (95% CI: 0.77-0.86), highlighting the need for improved RDTs or testing protocols^37^. Some studies highlight challenges with low specificity, indicating high false-positive rates that could lead to unnecessary treatments and strain on healthcare resources. For instance, one study reported high sensitivity (0.97, 95% CI: 0.93-0.99) but low specificity (0.39, 95% CI: 0.33-0.45), suggesting that while the RDTs are effective at detecting true malaria cases, they also falsely identify many non-malarial cases as positive^34^. This could overwhelm healthcare systems with false-positive results, leading to the inappropriate use of antimalarial medications and unnecessary healthcare costs.

Conversely, some studies report perfect specificity (1.00) but varying sensitivity^40–42^. While this indicates that the tests are excellent at ruling out non-malarial cases, the variable sensitivity suggests that they may miss some true malaria cases, which could lead to underdiagnosis and untreated infections. Studies from Ethiopia, Senegal, and Uganda consistently report high diagnostic accuracy, suggesting effective malaria detection in these areas^30,33^. Conversely, studies from Nigeria show lower sensitivity and specificity, indicating potential challenges in maintaining consistent diagnostic accuracy^34,35^. Factors such as local parasite diversity, environmental conditions, and test quality might contribute to this variability. In Uganda, some studies show high specificity (0.97 and 1.00, respectively) but varying sensitivity, reflecting generally good performance but highlighting areas for improvement in sensitivity^36,43^.

### Diagnostic Performance Based on Sample Type

The choice of sample type plays a crucial role in the reported diagnostic performance of RDTs. The studies analyzed provide insights into how different sample types, such as venous blood, capillary blood, dried blood spots, and peripheral blood, influence the sensitivity and specificity of RDTs in detecting malaria (Figure 7A and B). Venous blood, a commonly used sample type, generally demonstrates high specificity but varying sensitivity. Some studies reported high sensitivity (0.99, 95% CI: 0.97-1.00 and 0.95, 95% CI: 0.92-0.97, respectively) and specificity (0.98, 95% CI: 0.93-1.00 and 0.97, 95% CI: 0.95-0.99, respectively), suggesting strong diagnostic performance^30,44^. However, other studies reported lower sensitivity (0.49, 95% CI: 0.40-0.59 and 0.75, 95% CI: 0.72-0.78, respectively), despite maintaining moderate to high specificity^37,45^.

**Figure 7:**
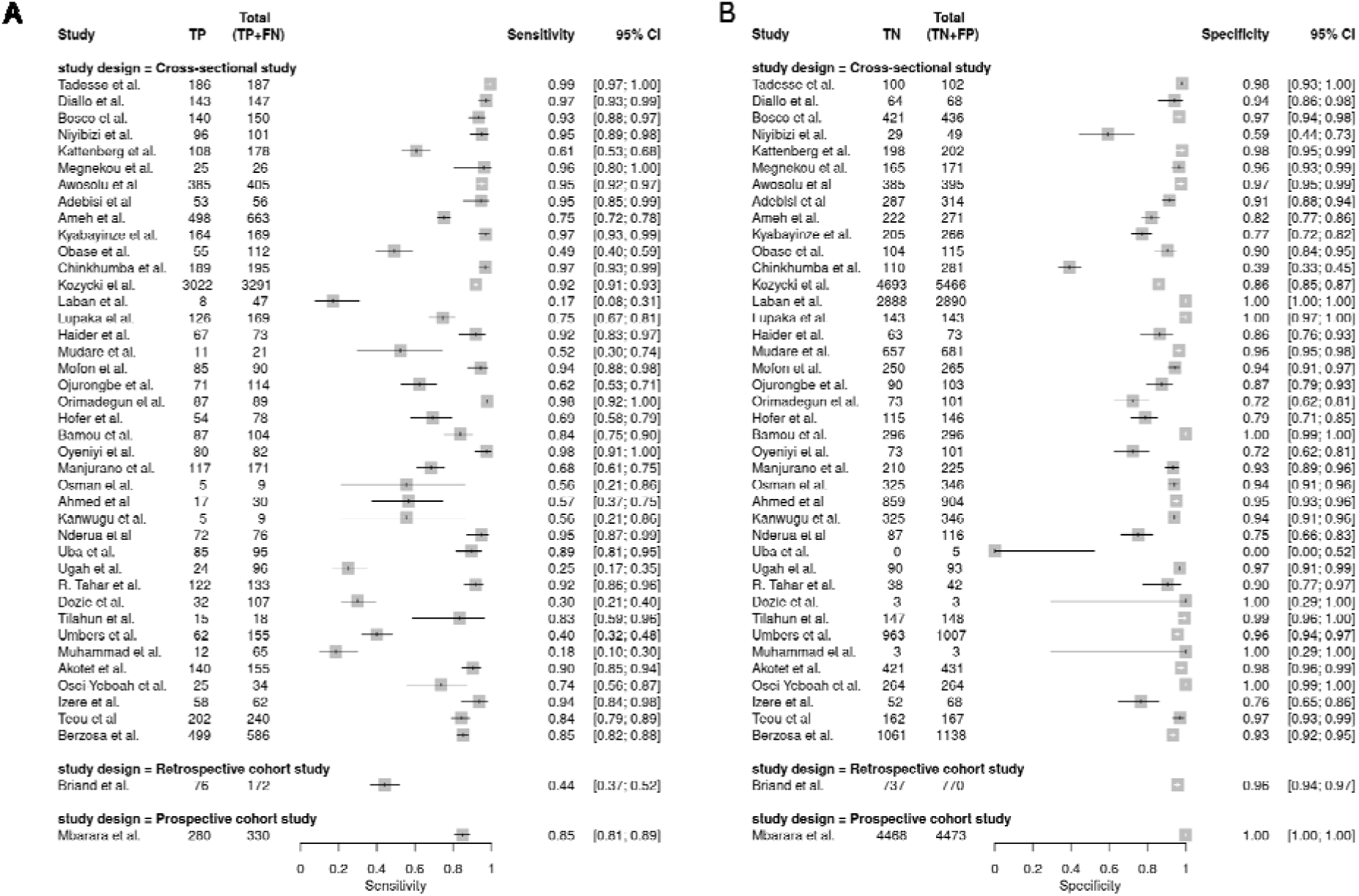
**(A)** Forest Plot of Sensitivity Estimates for Rapid Diagnostic Tests Based on Different Sample Types (B) Forest Plot of Specificity Estimates for Rapid Diagnostic Tests Based on Different Sample Types

Capillary blood, often collected via finger-prick, is another widely used sample type, particularly for its ease of collection. This sample type showed generally high diagnostic accuracy in studies, with sensitivities of 0.97 (95% CI: 0.93-0.99), 0.97 (95% CI: 0.93-0.99), 83.3(76.7–88.1) and specificities of 0.94 (95% CI: 0.86-0.98), 0.39 (95% CI: 0.33-0.45), 99.3(96.7–99.9), respectively^31,34,46^. However, some studies reported lower sensitivity and specificity, such as one reporting a sensitivity of 0.17 (95% CI: 0.08-0.31) and specificity of 1.00 (95% CI: 1.00-1.00)^36^.

Peripheral blood was evaluated in fewer studies but consistently showed strong diagnostic performance. One study reported a high sensitivity (0.96, 95% CI: 0.80-1.00) and specificity (0.96, 95% CI: 0.93-0.99)^47^. These findings suggest that peripheral blood, though less commonly used, offers a robust alternative for malaria diagnosis. The diagnostic performance of RDTs varies not only with sample type but also with the study design. Cross-sectional studies typically provide valuable snapshots of RDT performance in specific contexts. These studies often report high diagnostic accuracy, reflecting controlled assessments of RDTs^30,31^.

### Diagnostic Performance Based on RDT Brand

Examining the sensitivity and specificity of various rapid diagnostic test (RDT) brands for malaria reveals significant patterns and insights into their diagnostic performance. This analysis focuses on the performance of RDT brands such as SD BIOLINE (HRP2), CareStart (HRP2 and pLDH), First Response (HRP2), Paracheck (HRP2), Alere (HRP2), NADAL (HRP2), and Acon (HRP2 and pLDH). These RDTs target different antigens, such as Histidine-Rich Protein 2 (HRP2), Plasmodium lactate dehydrogenase (pLDH), and a combination of both, which influence their diagnostic performance (**Figure 8A and B**).

**Figure 8:**
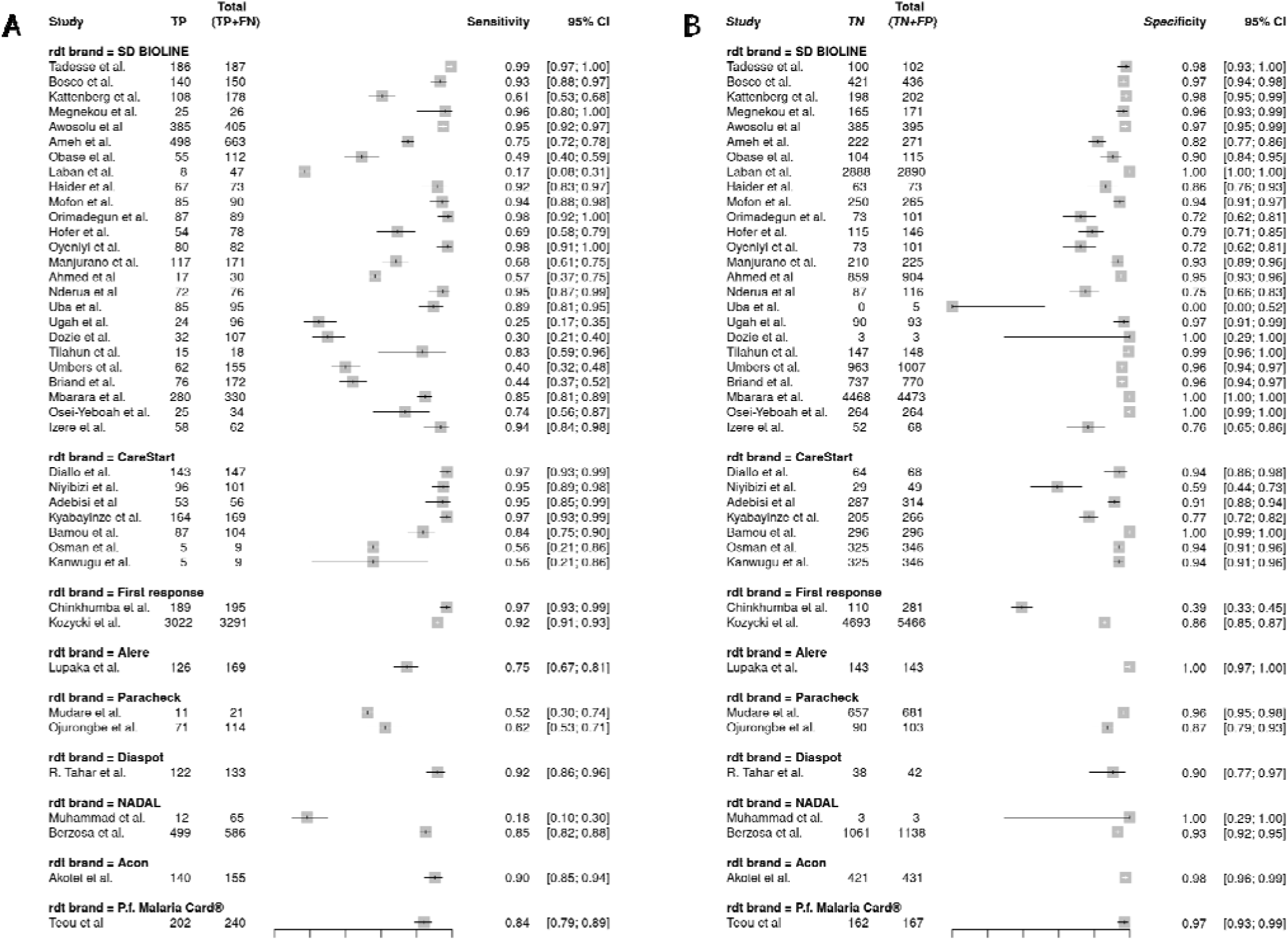
(A) Forest Plot of Sensitivity Estimates for Various Rapid Diagnostic Test Brands (B) Forest Plots of Specificity Estimates for Various Rapid Diagnostic Test Brands for Malaria

#### SD Bioline

SD Bioline is the most frequently used RDT brand in the studies, showing a wide range of sensitivity and specificity values. One study reported very high sensitivity (0.99, 95% CI: 0.97-1.00) and specificity (0.98, 95% CI: 0.93-1.00), ^30^. Other studies also showed high sensitivity (0.93, 95% CI: 0.88-0.97, and 0.95, 95% CI: 0.92-0.97, respectively) and specificity (0.97, 95% CI: 0.94-0.98, and 0.98, 95% CI: 0.95-0.99, respectively)^36,44^. However, some studies reported lower sensitivity (0.61, 95% CI: 0.53-0.68) despite high specificity (0.98, 95% CI: 0.95-0.99), suggesting potential issues with test accuracy in certain settings^39^. Another study showed high sensitivity (0.97, 95% CI: 0.93-0.99) but low specificity (0.39, 95% CI: 0.33-0.45), indicating a high rate of false positive^34^. These results highlight the variability in SD Bioline’s performance, which may influence regional factors, test administration, and parasite density.

#### CareStart

CareStart also demonstrates high diagnostic accuracy in many studies. One study reported a sensitivity of 0.97 (95% CI: 0.93-0.99) and a specificity of 0.94 (95% CI: 0.86-0.98)^31^, while another showed a sensitivity of 0.95 (95% CI: 0.85-0.99) and a specificity of 0.91 (95% CI: 0.88-0.94)^48^. Studies also reflected high specificity (1.00, 95% CI: 0.99-1.00, and 0.91, 95% CI: 0.88-0.94, respectively) with varying sensitivity (0.84, 95% CI: 0.75-0.90, and 0.95, 95% CI: 0.85-0.99, respectively)^40,48^. Another study reported perfect sensitivity (1.00, 95% CI: 0.97-1.00) and high specificity (0.94, 95% CI: 0.91-0.96), underscoring CareStart’s reliability^33^. However, some studies showed lower sensitivity (0.56, 95% CI: 0.21-0.86) and specificity (0.94, 95% CI: 0.91-0.96), indicating potential challenges in maintaining high diagnostic performance consistently^49,50^.

#### First Response

The First Response shows mixed results. A study reported high sensitivity (0.97, 95% CI: 0.93-0.99) but very low specificity (0.39, 95% CI: 0.33-0.45), suggesting a high false-positive rate^34^. A sensitivity of 0.92 (95% CI: 0.91-0.93) and a specificity of 0.86 (95% CI: 0.85-0.87) was recorded in another study, indicating moderate performance^33^. Another study reported a sensitivity of 0.97 (95% CI: 0.93-0.99) and specificity of 0.77 (95% CI: 0.72-0.82), reflecting reliable detection^51^. These mixed outcomes highlight the need to evaluate First Response RDTs further to improve their accuracy and reduce false positives.

#### Paracheck

Paracheck also demonstrates variable performance. One study reported a sensitivity of 0.52 (95% CI: 0.30-0.74) and a specificity of 0.97 (95% CI: 0.95-0.98), indicating moderate diagnostic accuracy^52^. Another study found a sensitivity of 0.62 (95% CI: 0.53-0.71) and specificity of 0.87 (95% CI: 0.79-0.93), reflecting similar moderate performance^53^. These results suggest that Paracheck RDTs may need enhancements to improve their sensitivity and overall diagnostic reliability.

#### Alere

Alere shows high specificity but variable sensitivity. One study reported a sensitivity of 0.75 (95% CI: 0.67-0.81) and perfect specificity (1.00, 95% CI: 0.97-1.00), indicating reliable exclusion of non-malarial cases but moderate sensitivity^35^. Another study showed lower sensitivity (0.44, 95% CI: 0.37-0.52) but high specificity (0.96, 95% CI: 0.94-0.97), reflecting similar patterns^54^. These results suggest that while Alere RDTs effectively avoid false positives, they may miss some true malaria cases, necessitating improvements in sensitivity.

#### NADAL

The study using NADAL RDTs reported very low sensitivity (0.18, 95% CI: 0.10-0.30) but perfect specificity (1.00, 95% CI: 0.29-1.00). This indicates significant challenges in accurately detecting malaria cases, highlighting the need for substantial improvements in the NADAL RDTs to enhance their diagnostic performance^55^.

#### Acon

Acon RDTs, as reported in another study, showed high sensitivity (0.90, 95% CI: 0.85-0.94) and specificity (0.98, 95% CI: 0.96-0.99), indicating reliable performance in detecting malaria cases^56^. This suggests that Acon RDTs effectively identify true malaria cases and avoid false positives, making them a valuable diagnostic tool.”

### Diagnostic Performance Based on Reference Standard Method

An analysis of the sensitivity and specificity of rapid diagnostic tests (RDTs) across multiple studies reveals the distinct impacts of microscopy and PCR as reference standard methods on diagnostic accuracy. The following discussion highlights the variations in diagnostic performance when using these two methods.

#### Microscopy

Microscopy remains a widely used reference standard in evaluating RDTs for malaria diagnosis, with generally high sensitivity and specificity reported across studies (**Figures 9 and 10**). For instance, one study reported a sensitivity of 0.99 (95% CI: 0.97-1.00) and a specificity of 0.98 (95% CI: 0.93-1.00), showcasing the reliability of microscopy in accurately identifying malaria cases^30^. Similarly, other studies demonstrated high diagnostic accuracy with sensitivities of 0.97 (95% CI: 0.93-0.99) and 0.93 (95% CI: 0.88-0.97), and specificities of 0.94 (95% CI: 0.86-0.98) and 0.97 (95% CI: 0.94-0.98), respectively^31,36^.

**Figure 9:**
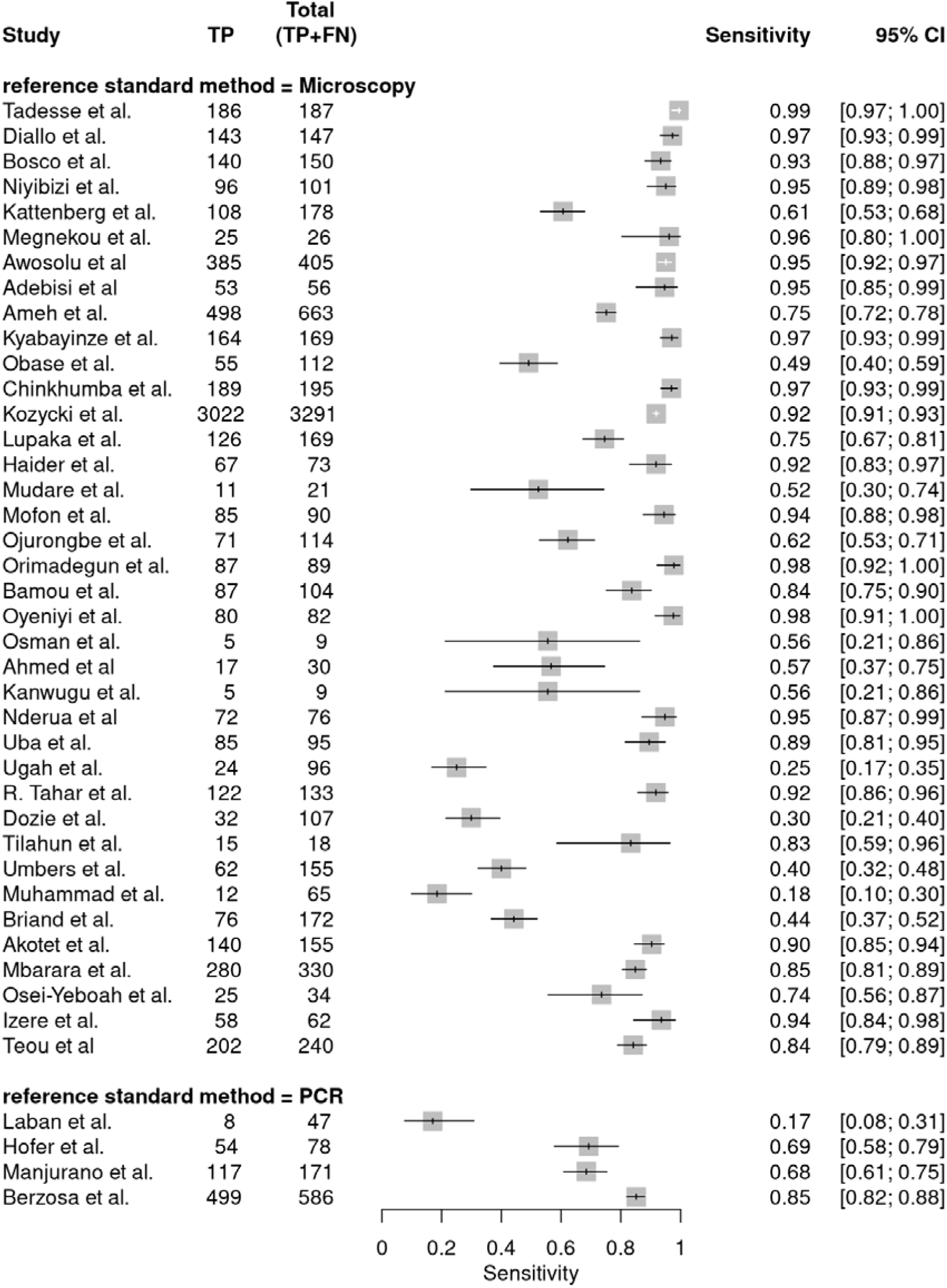
Forest Plot of Sensitivity Estimates for Rapid Diagnostic Tests Using Microscopy and PCR as Reference Standards.

**Figure 10:**
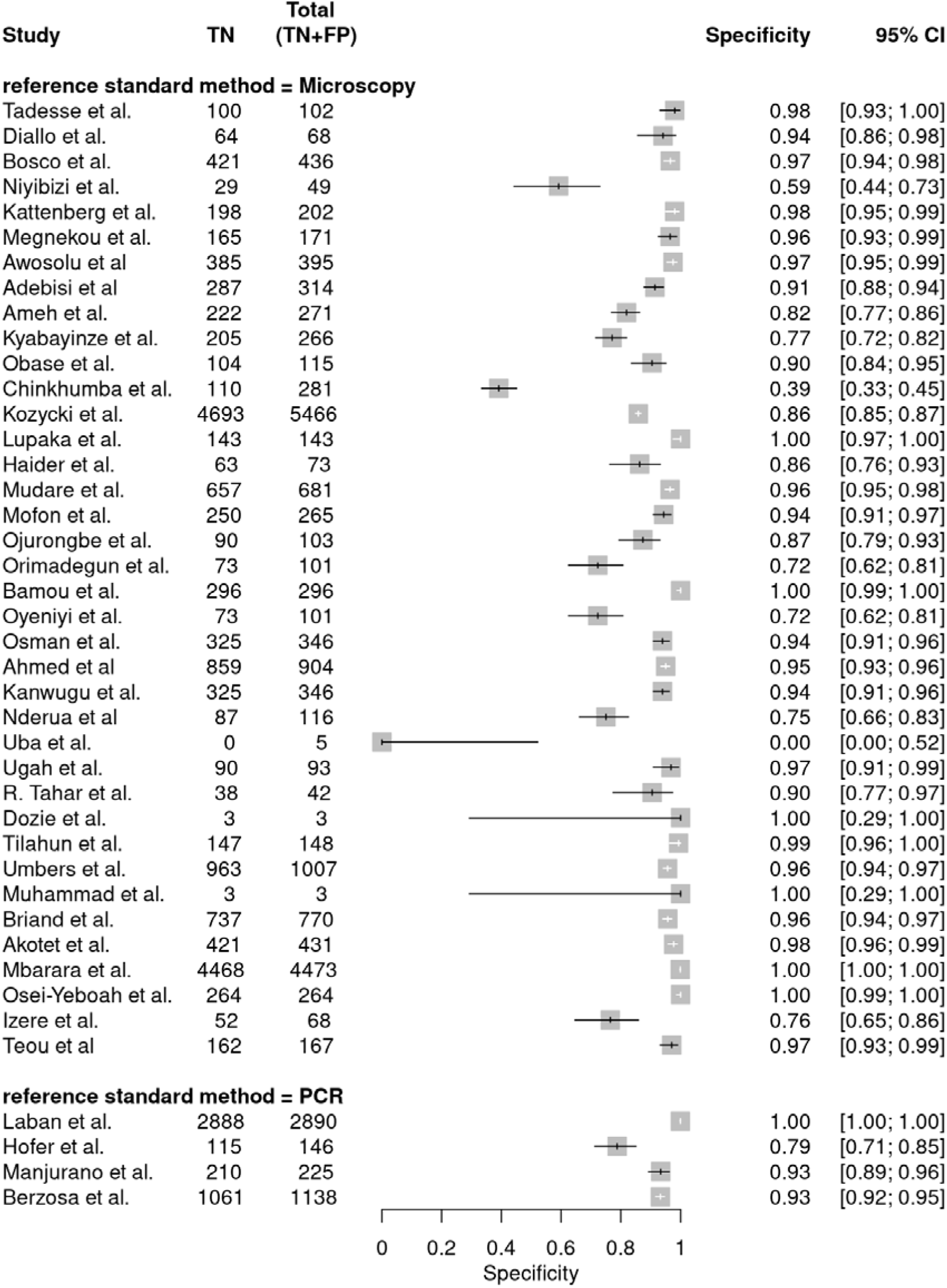
Forest Plot of Specificity Estimates for Rapid Diagnostic Tests Using Microscopy and PCR as Reference Standards.

However, some studies presented lower sensitivity, such as one reporting a sensitivity of 0.61 (95% CI: 0.53-0.68) despite a high specificity of 0.98 (95% CI: 0.95-0.99)^39^This suggests that while microscopy is generally reliable, sensitivity may be limited, particularly in specific settings or lower parasite densities.

#### Polymerase Chain Reaction (PCR)

PCR, particularly nested PCR (nPCR), serves as another critical reference standard for assessing the diagnostic performance of RDTs. The sensitivity and specificity of RDTs evaluated against PCR vary significantly (Figures 9 and 10). For example, one study reported a very low sensitivity of 0.17 (95% CI: 0.08-0.31) but perfect specificity (1.00, 95% CI: 1.00-1.00), indicating that while PCR is highly specific, its sensitivity may be compromised under certain conditions, possibly due to the low detection threshold required for some cases^32^. On the other hand, other studies demonstrated more balanced performance with sensitivities of 0.69 (95% CI: 0.58-0.79) and 0.85 (95% CI: 0.82-0.88), and specificities of 0.79 (95% CI: 0.71-0.85) and 0.93 (95% CI: 0.92-0.95), respectively^57,58^. These results highlight the variability in diagnostic performance when using PCR, which, while precise in excluding non-malarial cases, may face challenges in consistently detecting all true positive cases, especially when compared to microscopy. The analysis indicates that while both microscopy and PCR offer high specificity, ensuring the accurate exclusion of non-malarial cases, their sensitivity can vary depending on several factors, including the population under study and the specific conditions of the diagnostic tests. Microscopy generally offers more consistent sensitivity with occasional limitations, whereas PCR provides high specificity but may struggle with sensitivity in some contexts.

### Comparison and Statistical Analysis of RDTs Performance Over Time

Studies that evaluated the performance of various RDTs targeting malaria, specifically HRP2, Pf-specific LDH, and pan-specific LDH antigens, across different days following initial antimalarial treatment showed varying results. HRP2 sensitivity was 100% on Day 3, decreasing to 66.7% by Day 42. Specificity for HRP2 started low at 17.3% on Day 3, increasing to 94.8% by Day 42^59^. HRP2 RDTs demonstrated consistently high sensitivity of 100% across all time points from Day 3 to Day 42. However, specificity varied significantly, starting at 100% on Days 3 and 7, dropping to 20% on Day 14, and gradually increasing to 92.7% by Day 42^60^. HRP2 RDTs maintained 100% sensitivity, but specificity was initially low at 0% on Day 3, improving to 39.44% by Day 28^60^. Pan pLDH RDTs demonstrated high sensitivity and specificity, with sensitivity at 100% on Days 0, 21, and 28, and specificity ranging from 73.97% on Day 3 to 96.87% by Day 28^61^. HRP2 (SD Bioline) RDTs showed consistent sensitivity at 100% from Day 3 to Day 28, with specificity starting low at 25% on Days 3 and 7, increasing slightly to 33% by Day 28^62^. HRP2 (CareStart) RDTs revealed a high initial sensitivity of 100% on Day 0, decreasing slightly to 91.1% by Day 28. Specificity started high at 97.5% on Day 1 but decreased to 75.8% by Day 28^63^. Another study assessed HRP2 (ParaSight-F) RDTs, showing stable sensitivity at 93.3% across all evaluated days (Day 0, Day 7, Day 15, and Day 28), although specificity data was not available^64^

Comparing the performance of HRP2 RDTs over time, sensitivity generally remained high (>90%) initially but showed a slight decrease in some studies. Specificity for HRP2 RDTs exhibited significant variability, generally increasing over time. For example, one study reported a decrease in sensitivity from 100% on Day 0 to 91.1% by Day 28^63^. Specificity for HRP2 RDTs exhibited significant variability, generally increasing over time. Another study reported an increase in specificity from 17.3% on Day 3 to 94.8% by Day 42^59^. In contrast, Pf-specific LDH and panspecific LDH RDTs demonstrated consistently high sensitivity and specificity, often exceeding 90%. One study reported specificity reaching 100% by Day 42^59^.

When comparing HRP2 RDTs to other RDTs, HRP2 tests showed high initial sensitivity but variable specificity, often starting low and improving over time. Non-HRP2 RDTs like pan pLDH maintained high sensitivity and specificity from the start. For instance, another study reported pan pLDH RDTs with specificity ranging from 73.97% on Day 3 to 96.87% by Day 28, with high sensitivity throughout^61^. Statistical analysis revealed that across studies, HRP2 RDTs typically maintained high sensitivity (>90%) initially but may show a slight decrease over time. Specificity for HRP2 RDTs varied significantly but generally improved over time. Non-HRP2 RDTs, such as pan pLDH, initially exhibited high and stable specificity. Specific trends indicated that non-HRP2 RDTs might provide more consistent performance regarding specificity than HRP2 RDTs.

## DISCUSSION

This systematic review and meta-analysis evaluate the diagnostic performance of malaria RDTs in sub-Saharan Africa, identify diagnostic gaps, and propose strategies for novel test development. This discussion revisits the study’s objectives and highlights key findings, limitations, and implications for future research and practice. The comprehensive assessment of RDTs across sub-Saharan Africa revealed significant sensitivity, specificity, and variability in predictive values. High diagnostic accuracy was observed in studies targeting the HRP2 antigen specific to *Plasmodium falciparum*. Several studies reported sensitivities and specificities above 90%, particularly in high-transmission settings. For instance, two studies reported high sensitivity and specificity, emphasizing the effectiveness of these tests in detecting malaria in regions with a high burden of the disease^30,31^. However, a considerable range of performance was evident, with some studies reporting much lower diagnostic accuracy, highlighting the challenges in applying RDTs uniformly across diverse epidemiological contexts^66^.

The predictive values of the RDTs, particularly the positive predictive value (PPV) and negative predictive value (NPV), varied depending on malaria prevalence. The PPV was generally higher in high-prevalence settings, reflecting a greater likelihood that positive results truly indicated malaria. Conversely, the NPV was more critical in low-prevalence settings, ensuring that negative results reliably ruled out the disease. These findings highlight the importance of interpreting RDT results within the local epidemiological context to avoid misdiagnosis and inappropriate treatment. The heterogeneity analysis of the included studies revealed significant variability in the diagnostic performance of malaria rapid diagnostic tests (RDTs). The variance in the logit-transformed sensitivity was calculated at 2.333, while the variance in logit-transformed specificity was 3.305, indicating notable differences in sensitivity and specificity estimates across the studies. The median odds ratio (MOR) for sensitivity was 4.292, and for specificity, it was 5.664, further highlighting the variability in the ability of RDTs to correctly identify true positive and true negative cases between different studies. The bivariate I² statistic was found to be 0.733 (73.3%), demonstrating that a substantial portion of the variability in sensitivity and specificity is due to heterogeneity rather than random chance. Additionally, the area of the 95% prediction ellipse was 0.47, suggesting a moderate dispersion of sensitivity and specificity estimates around the mean.

The review identified several diagnostic gaps and limitations in current RDTs, particularly concerning their sensitivity to low-density parasitemia, the persistence of antigens leading to false positives, and cross-reactivity with other infections. A major limitation was the reduced sensitivity of RDTs in detecting low-density infections, especially in low-transmission settings. This was particularly pronounced in regions where asymptomatic or low-density infections are prevalent, as reported in certain studies^36^. The inability of RDTs to accurately detect these infections may contribute to underdiagnosis and ongoing transmission, highlighting a critical area for improvement in malaria diagnostics. Another significant challenge was the persistence of HRP2 antigenemia after treatment, which often led to false-positive results^30,31^. This phenomenon complicates the interpretation of RDTs in post-treatment scenarios, as persistent antigens can be mistaken for ongoing infection.

The deletion of the *pfhrp2* and *pfhrp3* genes has been frequently cited as a significant limitation in the effectiveness of HRP2-based RDTs. These can result in false-negative results, thereby reducing the utility of these tests in certain populations^36,65^. This issue is particularly pressing in areas where gene deletions are prevalent, underscoring the need for alternative diagnostic approaches.Some studies emphasize the need for alternative diagnostic targets or multi-antigen RDTs to address this limitation and improve post-treatment diagnostic accuracy^31,67^. Cross-reactivity with other febrile illnesses was also noted as a significant concern. In regions where other infections are common, this issue can lead to false-positive results, thereby reducing the specificity of RDTs. Addressing crossreactivity is essential to enhance the reliability of RDTs in diverse epidemiological settings^36^.

The performance of RDTs was influenced by geographical, epidemiological, and operational factors, necessitating the possibility of region-specific diagnostic strategies. Significant differences were observed in RDT performance across different regions. For instance, studies from Ethiopia and Uganda generally reported high diagnostic accuracy, likely due to effective malaria detection strategies and well-established diagnostic practices^30,36^. Conversely, studies from Nigeria and Malawi showed lower performance, underscoring the need for tailored diagnostic approaches that consider local conditions, including parasite diversity and transmission intensity. The diversity of *Plasmodium* species and varying transmission intensities also played a crucial role in influencing RDT performance. High-transmission areas generally reported better RDT performance, while regions with diverse *Plasmodium* species or lower transmission intensities faced challenges in maintaining high diagnostic accuracy. These findings underscore the importance of adapting diagnostic strategies to local epidemiological conditions and ensuring that RDTs are appropriately validated in specific regions.

Operational factors, such as the variability in test performance among health workers, further highlighted the need for robust training and quality assurance measures. Ensuring standardized test administration and handling procedures is crucial for obtaining reliable diagnostic outcomes across different settings, as highlighted by one study^34^. The genetic diversity of *Plasmodium* populations and polymorphisms, particularly in the HRP2 gene, can significantly impact the diagnostic accuracy of RDTs. Gene deletions, such as *pfhrp2/3* deletion, could be a major factor leading to falsenegative results, particularly in regions where these deletions are prevalent. There is a need for regular surveillance of parasite populations to monitor the prevalence of these polymorphisms and adapt diagnostic strategies accordingly. Moreover, the variability in antigen expression and genetic polymorphisms among parasite populations necessitates continuous monitoring and adaptation of diagnostic tools to ensure they remain effective against evolving parasite populations. This ongoing adaptation is crucial for maintaining the relevance and effectiveness of RDTs in different settings.

The importance of developing novel diagnostic tests or improving existing ones by leveraging emerging technologies and multi-platform approaches cannot be overemphasized. Advanced diagnostic technologies, such as next-generation sequencing and molecular diagnostics, offer promising avenues for enhancing the accuracy and reliability of malaria diagnostic tools. These technologies can provide detailed insights into parasite genetics, aiding in identifying novel diagnostic targets and developing more robust diagnostic platforms. Integrating multi-platform approaches, combining RDTs with other diagnostic methods like PCR and microscopy, was recommended to enhance overall diagnostic accuracy. Such approaches can leverage the strengths of different diagnostic methods, providing a more comprehensive solution to malaria diagnosis in sub-Saharan Africa.

Developing diagnostic tests tailored to the specific requirements of sub-Saharan Africa, considering local epidemiological conditions and healthcare infrastructure, was emphasized. This includes addressing issues like the erratic supply of test kits and ensuring consistent quality control across different RDT brands. The synthesis of studies evaluating the performance of various rapid diagnostic tests (RDTs) targeting malaria antigens, specifically HRP2 and Pf-specific LDH & panspecific LDH, reveals key insights into their effectiveness over time following antimalarial treatment. Overall, HRP2 RDTs demonstrate high initial sensitivity but exhibit variability in specificity, which tends to improve as the days post-treatment increase. In contrast, non-HRP2 RDTs, such as Pf-specific LDH and pan-specific LDH, maintain consistently high sensitivity and specificity throughout the evaluation period.

One study provided a comprehensive analysis of both HRP2 and Pf-specific LDH & pan-specific LDH RDTs. Their findings indicate that while HRP2 RDTs start with high sensitivity, specificity is initially low but improves significantly over time^59^. This trend is consistent with another study, which also found high sensitivity for HRP2 RDTs but noted significant fluctuations in specificity, particularly low specificity at the intermediate time points (e.g., Day 14)^60^. These variations in specificity could be attributed to the persistence of HRP2 antigenemia, which can remain detectable even after parasite clearance, leading to false-positive results.

The differences between HRP2 and pan pLDH RDTs were elucidated in a study highlighting the superior specificity of pan pLDH RDTs from the onset, in contrast to the initially low specificity of HRP2 RDTs. This suggests that pan pLDH RDTs may be more reliable in distinguishing between ongoing infection and post-treatment antigen presence^61^. Pan pLDH RDTs’ consistent performance in terms of sensitivity and specificity makes them a valuable tool for monitoring treatment efficacy, as evidenced by their stable high specificity and sensitivity across various time points.

Two studies focused on the HRP2 antigen, with both confirming high sensitivity but reporting varying specificity results^62,63^. One study observed consistently low specificity for HRP2 (SD Bioline) RDTs, which did not improve significantly over time^62^. Conversely, another study noted a decline in specificity over time for HRP2 (CareStart) RDTs, though initial specificity was high^63^. These findings underscore the need for careful interpretation of HRP2 RDT results, especially in the days immediately following treatment.

A longer-term perspective was provided in a study showing stable sensitivity for HRP2 (ParaSight-F) RDTs over a 28-day period, though specificity data was not available^64^. Their results align with the overall trend of high sensitivity for HRP2 RDTs but highlight the gap in data regarding specificity, which is crucial for accurate diagnosis and monitoring.

The overall analysis indicates that while HRP2 RDTs are highly sensitive initially, their specificity is often compromised shortly after treatment, leading to potential false positives. Non-HRP2 RDTs like pan pLDH offer a more consistent and reliable performance in both sensitivity and specificity, making them a preferable option for post-treatment monitoring. The variability in HRP2 RDT specificity suggests that their use should be complemented with other diagnostic methods or RDTs to ensure an accurate assessment of treatment efficacy.

### Conclusion

This systematic review and meta-analysis provided a comprehensive evaluation of malaria RDTs in sub-Saharan Africa, identifying key diagnostic gaps and proposing strategies for improvement. The findings underscore the importance of context-specific diagnostic approaches, continuous monitoring of test performance, and the development of novel diagnostic tools to enhance malaria detection and management in the region. Addressing the identified limitations and leveraging emerging technologies will be crucial for improving malaria diagnosis and ultimately reducing the disease burden in sub-Saharan Africa. While HRP2 RDTs demonstrate high initial sensitivity, their specificity is often compromised, particularly in the immediate post-treatment period. Non-HRP2 RDTs, such as pan pLDH, offer more stable and reliable diagnostic performance, making them a valuable tool for post-treatment monitoring. Future research should focus on identifying novel antigens and exploring the integration of multiple RDTs to further enhance diagnostic accuracy and reliability.

## METHODS

This systematic review and meta-analysis adhered to the PRISMA (Preferred Reporting Items for Systematic Reviews and Meta-Analyses) guidelines and is registered in PROSPERO with the identifier CRD42024551491. We conducted a comprehensive literature search via PubMed/MEDLINE and Google Scholar using keywords such as ‘Rapid Diagnostic Tests,’ ‘RDTs,’ ‘Malaria,’ ‘Plasmodium’, ‘Plasmodium species,’ ‘Sensitivity,’ ‘Specificity,’ ‘Diagnostic Gaps,’ and additional terms including ‘Antigen detection,’ ‘PCR,’ ‘Microscopy,’ ‘Diagnostic accuracy,’ ‘Malaria diagnostics,’ and ‘Test performance,’. We applied Medical Subject Headings (MeSH) terms, Boolean operators, and filters to refine the search.

### Study Selection and Data Extraction

Eligible studies included those that evaluated the diagnostic accuracy of malaria RDTs using microscopy or PCR as the reference standard and reported data sufficient to calculate sensitivity and specificity. Eligible studies encompassed original research articles, reviews, and conference proceedings published in English. The search strategies were tailored for each database, and the reference lists of included studies were manually checked for additional relevant citations. We used Rayyan (https://new.rayyan.ai/), an online systematic review tool, to facilitate the screening and selection. Two reviewers independently screened titles and abstracts to determine eligibility. Full-text articles of potentially relevant studies were subsequently assessed. Data extraction was conducted using a standardized form to ensure the collection of comprehensive and consistent information across studies. Extracted data included study characteristics (such as author(s), year of publication, study design, setting, sample size, study period, and season), population characteristics (age range, sex distribution, and specific population details), and details of the RDTs (brand, type, target antigen, Plasmodium species targeted, and sample type) (Supplementary Table S1). The types of RDTs were classified as Type I: HRP2-based RDTs (specifically target the Histidine-Rich Protein 2 (HRP2) antigen, which is specific to *Plasmodium falciparum),* Type II: HRP2 and aldolase RDTs (target both HRP2 and the aldolase antigen). Type III: Combined HRP2 and pLDH RDTs (detect both HRP2 and Plasmodium lactate dehydrogenase (pLDH, a biomarker for malaria infections). Type IV: Pf-specific LDH and panspecific LDH RDTs (target Plasmodium falciparum-specific LDH and pan-specific LDH antigens common to all malaria species). Additionally, performance metrics of the RDTs, including sensitivity, specificity, positive predictive value, and negative predictive value, along with the risk of bias assessments and applicability concerns, were also extracted (**Supplementary Tables S2, S3, and S4**). Information on reference standards reported limitations, and sample types were included in the studies (see Supplementary Table S5). Additionally, we documented the reference standard methods used in the studies and extracted diagnostic performance data, including the number of true positives, false positives, true negatives, and false negatives for both *P. falciparum* and *non-P. falciparum* species. We calculated sensitivity, specificity, positive predictive value (PPV), and negative predictive value (NPV) for each study. Other relevant information, such as parasite density, reported limitations, geographical region, and transmission setting, was also gathered.

### Quality Assessment and Data Synthesis

To evaluate the risk of bias and methodological quality of the included studies, we used the QUADAS-2 (Quality Assessment of Diagnostic Accuracy Studies 2) tool and RobVis ^29^ to produce a summary and graph of the risk of bias. This tool assesses studies across four domains: patient selection, index test, reference standard, and flow and timing.

#### Patient Selection

We described the methods of patient selection, including any prior testing, presentation details, the intended use of the index test, and the study setting. Signaling questions included whether a consecutive or random sample of patients was enrolled, whether a case-control design was avoided, and whether inappropriate exclusions were avoided. The risk of bias was evaluated based on whether the selection of patients could have introduced bias. Applicability concerns were assessed based on whether the included patients matched the review question.

#### Index Test

We described how the index test was conducted and interpreted. Signaling questions included whether the results of the index test were interpreted without knowledge of the reference standard results (blinding) and whether all patients underwent the same version of the index test. The risk of bias was evaluated based on whether the administration or interpretation of the index test could have introduced bias. Applicability concerns were assessed based on whether the index test, the way it was conducted, or the way it was interpreted aligned with the objectives of the review.

#### Reference Standard

We described the reference standard and how it was conducted and interpreted. Signaling questions included whether the reference standard correctly classified the target condition, whether the reference standard results were interpreted without knowledge of the index test results, and whether all patients received the reference standard. The risk of bias was evaluated based on whether the conduct (i.e., the procedures followed in administering and interpreting the reference standard test) or the interpretation of the reference standard could have introduced bias. Applicability concerns were assessed based on whether the target condition defined by the reference standard matched the review question.

#### Flow and Timing

We described any patients who did not receive the index test(s) and/or reference standard, were excluded from the 2퀇2 table, the time interval between the index test(s) and reference standard, and any interventions. Signaling questions included whether there was an appropriate interval between index test(s) and reference standard, whether all patients received a reference standard, whether all patients received the same reference standard, and whether all patients were included in the analysis. The risk of bias was evaluated based on whether the patient flow could have introduced bias.

For data synthesis, we utilized the MetaDTA (https://crsu.shinyapps.io/MetaDTA/) online application for statistical analysis. This tool enabled us to perform a robust analysis of diagnostic test accuracy by providing advanced statistical methods for pooling and comparing diagnostic performance metrics. A random effects model was employed to account for the variability between studies included in the meta-analysis. This model is particularly suitable for meta-analyses involving studies with heterogeneous designs, populations, and settings, ensuring that the pooled estimates of sensitivity and specificity are robust and generalizable. We employed visual inspection and statistical methods to further assess the heterogeneity among the included studies. We calculated the variance in the logit-transformed sensitivity and specificity to quantify the variability in diagnostic performance across studies. The variance in logit sensitivity and specificity was computed to capture the extent of differences in these estimates among the studies. Additionally, we calculated the median odds ratio (MOR) for sensitivity and specificity to measure the variability in the odds of correctly identifying true positive and true negative cases between different studies. The MOR indicates the relative difference in diagnostic accuracy across the included studies. We used the bivariate I² statistic to quantify the overall heterogeneity, representing the proportion of total variability in sensitivity and specificity attributable to heterogeneity rather than chance. I² value close to 1 indicates a high level of heterogeneity. We assessed the dispersion of sensitivity and specificity estimates using the area of the 95% prediction ellipse derived from the summary ROC (SROC) curve. This measure helps visualize the estimates spread around the mean diagnostic accuracy. These statistical measures were selected to provide a comprehensive understanding of the heterogeneity in the diagnostic accuracy of malaria RDTs across different settings and study populations.

## Author contributions

F.D.O. conceptualized the study, designed the systematic review and meta-analysis methodology, screened papers, extracted data, and led the writing of the original draft. A.O.A. contributed to reviewing and editing the manuscript, wrote the abstract, and provided critical feedback and necessary corrections. O.I. conducted the literature search, performed screening, extracted data, validated data accuracy, and reviewed the manuscript. OOO, OAO, and L.O.E coordinated the research activities, ensured adherence to timelines, and contributed to reviewing and editing the manuscript. B.T. and O.O. contributed to the review and final editing. O.O. conceptualized, supervised the research team, and provided mentorship.

## Data availability statement

The datasets generated and/or analyzed during the current study are available in the Mendeley Data repository, doi: 10.17632/2wbv7yfzbr.1.

