## Supplementary file for "Performance and Challenges of Malaria Rapid Diagnostic Tests in Endemic Regions of Africa"

TABLE S1: Diagnostic Performance of Various Studies on Malaria Rapid Diagnostic Tests

| study | Study Design | Sens | Spec |
| --- | --- | --- | --- |
| Tadesse et al. | Diagnostic Performance Evaluation | 0.995 | 0.98 |
| Diallo et al. | Diagnostic Accuracy Study | 0.973 | 0.941 |
| Bosco et al. | Cross-sectional Study | 0.933 | 0.966 |
| Kattenberg et al. | Cross-Sectional Study | 0.607 | 0.98 |
| Alareqi et al | Cross-sectional | 0.963 | 0.561 |
| Awosolu et al | Cross-sectional | 0.951 | 0.975 |
| Adebisi_2018 et al | Cross-sectional | 0.946 | 0.914 |
| Ameh et al. | Cross-sectional | 0.751 | 0.819 |
| Azazy et al. | Cross-sectional | 1 | 0.973 |
| Obase et al. | Cross-sectional | 0.491 | 0.904 |
| Chinkhumba et al. | Cross-sectional | 0.969 | 0.391 |
| Kozycki et al. | Cross-sectional | 0.918 | 0.859 |
| Laban et al. | Cross-sectional | 0.17 | 0.999 |
| Lupaka et al. | Cross-sectional | 0.746 | 1 |
| Haider et al. | Cross-sectional | 0.918 | 0.863 |
| Mudare et al. | Cross-sectional | 0.524 | 0.965 |
| X et al. | Cross-sectional | 0.944 | 0.943 |
| Ojurongbe et al. | Cross-sectional | 0.623 | 0.874 |
| Orimadegun et al. | Cross-sectional | 0.978 | 0.723 |
| Y et al. | Cross-sectional | 0.909 | 0.929 |
| Bamou et al. | Cross-sectional | 0.837 | 1 |
| Adebisi_2022 et al. | Cross-sectional | 0.946 | 0.914 |
| Oyeniyi et al. | Cross-sectional | 0.976 | 0.723 |
| Z et al. | Cross-sectional | 1 | 0.938 |
| Osman et al. | Cross-sectional | 0.556 | 0.939 |
| Ahmed et al | Cross-sectional | 0.567 | 0.95 |
| Kanwugu et al. | Cross-sectional | 0.556 | 0.939 |
| Nderua et al (microscopy) | Cross-sectional | 0.947 | 0.75 |
| Uba et al. | Cross-sectional | 0.895 | 0 |
| Ugah et al. | Cross-sectional | 0.25 | 0.968 |
| Dozie et al. | Cross-sectional | 0.299 | 1 |
| Tilahun et al. | Cross-sectional | 0.833 | 0.993 |
| Umbers et al. | Cross-sectional | 0.4 | 0.956 |
| Zainab Sabo Muhammad et al. | Cross-sectional | 0.185 | 1 |
| Briand et al. | Retrospective cohort study | 0.442 | 0.957 |
| Bouyou Akotet et al. | Cross-sectional | 0.903 | 0.977 |
| Grandesso_Mbarara et al. | Prospective cohort study | 0.848 | 0.999 |
| James Osei-Yeboah et al. | Cross-sectional study | 0.735 | 1 |

TABLE S2: **Diagnostic Performance and Plasmodium Species Targeted in Various Malaria Studies**

| study | Plasmodium Species Targeted | Sens | Spec |
| --- | --- | --- | --- |
| Tadesse et al. | P. falciparum, non-P. falciparum | 0.995 | 0.98 |
| Diallo et al. | P. falciparum | 0.973 | 0.941 |
| Bosco et al. | P. falciparum | 0.933 | 0.966 |
| Kattenberg et al. | P. falciparum, P. vivax | 0.607 | 0.98 |
| Alareqi et al | Plasmodium falciparum | 0.963 | 0.561 |
| Awosolu et al | Plasmodium falciparum | 0.951 | 0.975 |
| Adebisi_2018 et al | Plasmodium falciparum | 0.946 | 0.914 |
| Ameh et al. | Plasmodium falciparum and Plasmodium vivax | 0.751 | 0.819 |
| Azazy et al. | Plasmodium falciparum | 1 | 0.973 |
| Obase et al. | Plasmodium falciparum and Plasmodium ovale | 0.491 | 0.904 |
| Chinkhumba et al. | Plasmodium falciparum | 0.969 | 0.391 |
| Kozycki et al. | Plasmodium falciparum | 0.918 | 0.859 |
| Laban et al. | Plasmodium falciparum, Plasmodium malariae | 0.17 | 0.999 |
| Lupaka et al. | Plasmodium falciparum | 0.746 | 1 |
| Haider et al. | P. falciparum and others | 0.918 | 0.863 |
| Mudare et al. | Plasmodium falciparum | 0.524 | 0.965 |
| X et al. | Plasmodium falciparum | 0.944 | 0.943 |
| Ojurongbe et al. | Plasmodium falciparum | 0.623 | 0.874 |
| Orimadegun et al. | Plasmodium falciparum | 0.978 | 0.723 |
| Y et al. | Plasmodium falciparum | 0.909 | 0.929 |
| Bamou et al. | Plasmodium falciparum | 0.837 | 1 |
| Adebisi_2022 et al. | Plasmodium falciparum | 0.946 | 0.914 |
| Oyeniyi et al. | Plasmodium falciparum | 0.976 | 0.723 |
| Z et al. | Plasmodium falciparum | 1 | 0.938 |
| Osman et al. | Plasmodium falciparum | 0.556 | 0.939 |
| Ahmed et al | Plasmodium falciparum | 0.567 | 0.95 |
| Kanwugu et al. | Plasmodium falciparum | 0.556 | 0.939 |
| Nderua et al (microscopy) | Plasmodium falciparum | 0.947 | 0.75 |
| Uba et al. | Plasmodium falciparum | 0.895 | 0 |
| Ugah et al. | Plasmodium falciparum | 0.25 | 0.968 |
| Dozie et al. | Plasmodium falciparum | 0.299 | 1 |
| Tilahun et al. | Plasmodium falciparum | 0.833 | 0.993 |
| Umbers et al. | Plasmodium falciparum | 0.4 | 0.956 |
| Zainab Sabo Muhammad et al. | Plasmodium falciparum, Plasmodium vivax, Plasmodium ovale, Plasmodium malariae | 0.185 | 1 |
| Briand et al. | Plasmodium falciparum | 0.442 | 0.957 |
| Bouyou Akotet et al. | Plasmodium falciparum, Plasmodium ovale, Plasmodium malariae | 0.903 | 0.977 |
| Grandesso_Mbarara et al. | Plasmodium falciparum | 0.848 | 0.999 |
| James Osei-Yeboah et al. | Plasmodium falciparum | 0.735 | 1 |

TABLE S3: Diagnostic Performance of Malaria Rapid Diagnostic Tests (RDTs) by Brand

| study | RDT Brand | Sens | Spec |
| --- | --- | --- | --- |
| Tadesse et al. | SD BIOLINE | 0.995 | 0.98 |
| Diallo et al. | CareStart | 0.973 | 0.941 |
| Bosco et al. | SD BIOLINE | 0.933 | 0.966 |
| Kattenberg et al. | SD BIOLINE | 0.607 | 0.98 |
| Alareqi et al | SD BIOLINE | 0.963 | 0.561 |
| Awosolu et al | SD BIOLINE | 0.951 | 0.975 |
| Adebisi_2018 et al | CareStart | 0.946 | 0.914 |
| Ameh et al. | SD BIOLINE | 0.751 | 0.819 |
| Azazy et al. | SD BIOLINE | 1 | 0.973 |
| Obase et al. | SD BIOLINE | 0.491 | 0.904 |
| Chinkhumba et al. | First response | 0.969 | 0.391 |
| Kozycki et al. | First response | 0.918 | 0.859 |
| Laban et al. | SD BIOLINE | 0.17 | 0.999 |
| Lupaka et al. | Alere | 0.746 | 1 |
| Haider et al. | SD BIOLINE | 0.918 | 0.863 |
| Mudare et al. | Paracheck | 0.524 | 0.965 |
| X et al. | SD BIOLINE | 0.944 | 0.943 |
| Ojurongbe et al. | Paracheck | 0.623 | 0.874 |
| Orimadegun et al. | SD BIOLINE | 0.978 | 0.723 |
| Y et al. | First response | 0.909 | 0.929 |
| Bamou et al. | CareStart | 0.837 | 1 |
| Adebisi_2022 et al. | CareStart | 0.946 | 0.914 |
| Oyeniyi et al. | SD BIOLINE | 0.976 | 0.723 |
| Z et al. | CareStart | 1 | 0.938 |
| Osman et al. | CareStart | 0.556 | 0.939 |
| Ahmed et al | SD BIOLINE | 0.567 | 0.95 |
| Kanwugu et al. | CareStart | 0.556 | 0.939 |
| Nderua et al (microscopy) | SD BIOLINE | 0.947 | 0.75 |
| Uba et al. | SD BIOLINE | 0.895 | 0 |
| Ugah et al. | SD BIOLINE | 0.25 | 0.968 |
| Dozie et al. | SD BIOLINE | 0.299 | 1 |
| Tilahun et al. | Not specified | 0.833 | 0.993 |
| Umbers et al. | SD BIOLINE | 0.4 | 0.956 |
| Zainab Sabo Muhammad et al. | NADAL | 0.185 | 1 |
| Briand et al. | Alere | 0.442 | 0.957 |
| Bouyou Akotet et al. | Acon | 0.903 | 0.977 |
| Grandesso_Mbarara et al. | SD BIOLINE | 0.848 | 0.999 |
| James Osei-Yeboah et al. | SD BIOLINE | 0.735 | 1 |

TABLE S4 Sensitivity and Specificity of Malaria Diagnostic Studies Using Various Reference Standard Methods

| Study | Reference Standard Method | Sens | Spec |
| --- | --- | --- | --- |
| Tadesse et al. | Microscopy of Giemsa-stained blood films | 0.995 | 0.98 |
| Diallo et al. | Microscopy, PCR | 0.973 | 0.941 |
| Bosco et al. | Microscopy, PCR | 0.933 | 0.966 |
| Kattenberg et al. | Microscopy, PCR | 0.607 | 0.98 |
| Alareqi et al | Nested Polymerase Chain Reaction (PCR) | 0.963 | 0.561 |
| Awosolu et al | Microscopy | 0.951 | 0.975 |
| Adebisi et al | Microscopy | 0.946 | 0.914 |
| Ameh et al. | Microscopy | 0.751 | 0.819 |
| Azazy et al. | Microscopy | 1 | 0.973 |
| Obase et al. | Microscopy, PCR, Composite Test | 0.491 | 0.904 |
| Chinkhumba et al. | Microscopy | 0.969 | 0.391 |
| Kozycki et al. | Microscopy | 0.918 | 0.859 |
| Laban et al. | Nested PCR | 0.17 | 0.999 |
| Lupaka et al. | Microscopy and PCR | 0.746 | 1 |
| Haider et al. | Microscopy | 0.918 | 0.863 |
| Mudare et al. | PCR, Microscopy | 0.524 | 0.965 |
| X et al. | Microscopy, PCR | 0.944 | 0.943 |
| Ojurongbe et al. | Microscopy, PCR | 0.623 | 0.874 |
| Orimadegun et al. | Microscopy, PCR | 0.978 | 0.723 |
| Y et al. | Microscopy, PCR | 0.909 | 0.929 |
| Bamou et al. | Microscopy | 0.837 | 1 |
| Adebisi et al. | Microscopy | 0.946 | 0.914 |
| Oyeniyi et al. | Microscopy | 0.976 | 0.723 |
| Z et al. | Microscopy | 1 | 0.938 |
| Osman et al. | Microscopy | 0.556 | 0.939 |
| Ahmed et al | Microscopy, PCR | 0.567 | 0.95 |
| Kanwugu et al. | Microscopy | 0.556 | 0.939 |
| Nderua et al (microscopy) | Microscopy, PCR | 0.947 | 0.75 |
| Uba et al. | Microscopy, nPCR | 0.895 | 0 |
| Ugah et al. | Microscopy, PCR | 0.25 | 0.968 |
| Dozie et al. | Microscopy | 0.299 | 1 |
| Tilahun et al. | Microscopy, RT-PCR | 0.833 | 0.993 |
| Umbers et al. | Microscopy, PCR | 0.4 | 0.956 |
| Zainab Sabo Muhammad et al. | Microscopy | 0.185 | 1 |
| Briand et al. | qPCR, Microscopy | 0.442 | 0.957 |
| Bouyou Akotet et al. | Microscopy | 0.903 | 0.977 |
| Grandesso_Mbarara et al. | Microscopy | 0.848 | 0.999 |
| James Osei-Yeboah et al. | Microscopy | 0.735 | 1 |

TABLE S5: Sensitivity and Specificity of Malaria Diagnostic Tests by Sample Type

| study | Sample Type | Sens | Spec |
| --- | --- | --- | --- |
| Tadesse et al. | Venous blood, finger-prick blood | 0.995 | 0.98 |
| Diallo et al. | Finger-prick, venous blood | 0.973 | 0.941 |
| Bosco et al. | Dried blood spots | 0.933 | 0.966 |
| Kattenberg et al. | Venous blood | 0.607 | 0.98 |
| Alareqi et al | Whole blood samples | 0.963 | 0.561 |
| Awosolu et al | Venous blood samples | 0.951 | 0.975 |
| Adebisi_2018 et al | Finger prick blood samples | 0.946 | 0.914 |
| Ameh et al. | EDTA blood samples | 0.751 | 0.819 |
| Azazy et al. | EDTA blood samples | 1 | 0.973 |
| Obase et al. | Mother venous and cord blood | 0.491 | 0.904 |
| Chinkhumba et al. | Finger prick blood samples | 0.969 | 0.391 |
| Kozycki et al. | Finger prick blood samples | 0.918 | 0.859 |
| Laban et al. | Finger prick blood samples | 0.17 | 0.999 |
| Lupaka et al. | Finger-prick blood | 0.746 | 1 |
| Haider et al. | Venous blood | 0.918 | 0.863 |
| Mudare et al. | Finger-prick blood | 0.524 | 0.965 |
| X et al. | Venous blood | 0.944 | 0.943 |
| Ojurongbe et al. | Venous blood | 0.623 | 0.874 |
| Orimadegun et al. | Finger-prick blood | 0.978 | 0.723 |
| Y et al. | Venous blood | 0.909 | 0.929 |
| Bamou et al. | Venous blood | 0.837 | 1 |
| Adebisi_2022 et al. | Finger-prick blood | 0.946 | 0.914 |
| Oyeniyi et al. | Venous blood | 0.976 | 0.723 |
| Z et al. | Venous blood | 1 | 0.938 |
| Osman et al. | Finger-prick blood | 0.556 | 0.939 |
| Ahmed et al | Venous blood | 0.567 | 0.95 |
| Kanwugu et al. | Finger-prick blood | 0.556 | 0.939 |
| Nderua et al (microscopy) | Venous blood | 0.947 | 0.75 |
| Uba et al. | Venous blood | 0.895 | 0 |
| Ugah et al. | Venous blood | 0.25 | 0.968 |
| Dozie et al. | Venous blood | 0.299 | 1 |
| Tilahun et al. | Dried blood spot (DBS) | 0.833 | 0.993 |
| Umbers et al. | Venous blood | 0.4 | 0.956 |
| Zainab Sabo Muhammad et al. | Finger-prick blood | 0.185 | 1 |
| Briand et al. | Venous blood | 0.442 | 0.957 |
| Bouyou Akotet et al. | Venous blood | 0.903 | 0.977 |
| Grandesso_Mbarara et al. | Venous blood | 0.848 | 0.999 |
| James Osei-Yeboah et al. | Blood sample | 0.735 | 1 |

TABLE S6: Sensitivity and Specificity of Malaria Diagnostic Studies with Limitations

| study | Limitations | Sens | Spec |
| --- | --- | --- | --- |
| Tadesse et al. | Persistence of HRP2 after treatment, variation in antigen concentration affecting detection | 0.995 | 0.98 |
| Diallo et al. | Persistence of HRP2 antigen after treatment, effect of heat/humidity on pLDH | 0.973 | 0.941 |
| Bosco et al. | pfhrp2/3 gene deletion, non-P. falciparum species, low-density infections | 0.933 | 0.966 |
| Kattenberg et al. | Antigen persistence, storage issues with ELISA, impact of gametocytes on HRP2 persistence | 0.607 | 0.98 |
| Alareqi et al | High false-positivity rate for RDT, low sensitivity of LM | 0.963 | 0.561 |
| Awosolu et al | High false positivity rate, low sensitivity for low parasite density | 0.951 | 0.975 |
| Adebisi_2018 et al | High false positivity rate, limited confirmation with PCR | 0.946 | 0.914 |
| Ameh et al. | High false negative rate, prozone effect for high parasite density | 0.751 | 0.819 |
| Azazy et al. | Not a field study, may overlook utility for sub-microscopic infections | 1 | 0.973 |
| Obase et al. | High false positives, PCR more sensitive but resource-intensive | 0.491 | 0.904 |
| Chinkhumba et al. | Low specificity, variable performance by health workers, need for robust quality assurance | 0.969 | 0.391 |
| Kozycki et al. | Low sensitivity of HRP2 RDT in low transmission settings, hrp2 deletions | 0.918 | 0.859 |
| Laban et al. | Low sensitivity to low-density infections, not detecting non-falciparum malaria | 0.17 | 0.999 |
| Lupaka et al. | False negatives due to low parasite density and possible HRP2/3 gene deletions | 0.746 | 1 |
| Haider et al. | Lower sensitivity and specificity of RDT compared to ELISA, ease of performing the test, rapid outcomes, and ability to differentiate between P. falciparum and P. vivax highlighted | 0.918 | 0.863 |
| Mudare et al. | Did not assess for pfhrp2/3 deletions, potential false positives due to persisting antigenaemia | 0.524 | 0.965 |
| X et al. | Persistence of HRP2 antigenemia could result in false positives | 0.944 | 0.943 |
| Ojurongbe et al. | Persistence of HRP2 antigenemia could result in false positives | 0.623 | 0.874 |
| Orimadegun et al. | False positives due to HRP2 antigen persistence, potential HRP2 gene deletions | 0.978 | 0.723 |
| Y et al. | Potential HRP2 gene deletions, cross-reactivity with other febrile illnesses | 0.909 | 0.929 |
| Bamou et al. | Low sensitivity at low parasite density, potential for false positives due to HRP2 persistence | 0.837 | 1 |
| Adebisi_2022 et al. | Erratic supply of RDT kits, non-availability of test kits, varying sensitivity reports in Nigeria | 0.946 | 0.914 |
| Oyeniyi et al. | Potential cross-reactivity with other infections, persistence of HRP2 antigen | 0.976 | 0.723 |
| Z et al. | Potential cross-reactivity with other febrile illnesses, persistence of HRP2 antigen | 1 | 0.938 |
| Osman et al. | Low sensitivity at low parasite density, potential false positives due to HRP2 persistence | 0.556 | 0.939 |
| Ahmed et al | Potential false positives due to persistence of HRP2, variable sensitivity among different brands | 0.567 | 0.95 |
| Kanwugu et al. | Low sensitivity at low parasite density, potential false positives due to HRP2 persistence | 0.556 | 0.939 |
| Nderua et al (microscopy) | Potential false positives due to persistence of HRP2, variable sensitivity among different brands | 0.947 | 0.75 |
| Uba et al. | Erratic supply of RDT kits, false positives due to persistence of HRP2 antigen, lower sensitivity in nPCR | 0.895 | 0 |
| Ugah et al. | Potential false positives due to HRP2 persistence, varying sensitivity among different brands | 0.25 | 0.968 |
| Dozie et al. | Lower sensitivity of RDT compared to microscopy, high specificity | 0.299 | 1 |
| Tilahun et al. | Variability in sensitivity and specificity among diagnostic methods, importance of using RT-PCR for accurate diagnosis among asymptomatic individuals | 0.833 | 0.993 |
| Umbers et al. | Potential false positives due to persistence of HRP2, variable sensitivity among different brands | 0.4 | 0.956 |
| Zainab Sabo Muhammad et al. | Low sensitivity of RDT compared to microscopy, high false negative rate for RDT, need for better training and handling of test kits | 0.185 | 1 |
| Briand et al. | Lower sensitivity of cRDT, higher false negatives for cRDT, importance of uRDT in detecting low-density infections, association with maternal anemia | 0.442 | 0.957 |
| Bouyou Akotet et al. | High false positive rate for Pf/Pan RDT, variability in sensitivity among different age groups, potential misinterpretation due to faint bands | 0.903 | 0.977 |
| Grandesso_Mbarara et al. | Lower specificity of HRP2 in high-transmission settings, long time to negativity for HRP2 tests, variable performance based on transmission setting and parasite density | 0.848 | 0.999 |
| James Osei-Yeboah et al. | Low sensitivity and specificity, RDT accuracy affected by product quality and user performance | 0.735 | 1 |

TABLE S7: Sensitivity and Specificity of Malaria Rapid Diagnostic Tests (RDTs) by Target Antigens and Days After Initial Treatment

| Author | Year | RDTs Target Antigens | Days After Initial Treatment | TP | TN | FP | FN | Sensitivity (%) | Specificity (%) |
| --- | --- | --- | --- | --- | --- | --- | --- | --- | --- |
| Houzé et al. | 2009 | HRP2 | Day 3 | 35 | 28 | 134 | 0 | 100 | 17.3 |
|  |  |  | Day 7 | 6 | 49 | 115 | 1 | 85.7 | 29.9 |
|  |  |  | Day 14 | 6 | 87 | 69 | 1 | 85.7 | 55.8 |
|  |  |  | Day 21 | 14 | 104 | 38 | 2 | 87.5 | 73.2 |
|  |  |  | Day 28 | 13 | 92 | 63 | 2 | 86.7 | 73.6 |
|  |  |  | Day 35 | 9 | 79 | 4 | 2 | 81.8 | 95.2 |
|  |  |  | Day 42 | 2 | 73 | 4 | 1 | 66.7 | 94.8 |
|  |  | Pf-specific LDH & pan-specific LDH | Day 3 | 28 | 141 | 21 | 7 | 80 | 87.1 |
|  |  |  | Day 7 | 5 | 151 | 13 | 2 | 71.4 | 92 |
|  |  |  | Day 14 | 5 | 150 | 6 | 2 | 71.4 | 96.1 |
|  |  |  | Day 21 | 16 | 137 | 5 | 0 | 100 | 96.5 |
|  |  |  | Day 28 | 13 | 122 | 3 | 2 | 86.7 | 97.6 |
|  |  |  | Day 35 | 9 | 81 | 2 | 2 | 81.8 | 97.6 |
|  |  |  | Day 42 | 2 | 77 | 0 | 1 | 66.7 | 100 |
| Aydin-Schmidt et al. | 2013 | HRP2 | Day 3 | 11 | 5 | 0 | 0 | 100 | 100 |
|  |  |  | Day 7 | 7 | 3 | 0 | 0 | 100 | 100 |
|  |  |  | Day 14 | 3 | 8 | 32 | 0 | 100 | 20 |
|  |  |  | Day 21 | 1 | 15 | 27 | 0 | 100 | 35.7 |
|  |  |  | Day 28 | 1 | 24 | 18 | 0 | 100 | 57.1 |
|  |  |  | Day 35 | 1 | 32 | 10 | 0 | 100 | 76.2 |
|  |  |  | Day 42 | 2 | 38 | 3 | 0 | 100 | 92.7 |
| Nyunt et al. | 2013 | HRP2 | Day 0 | 77 | NR | NR | 0 | 100 | NR |
|  |  |  | Day 3 | 77 | 0 | 59 | 0 | 100 | 0 |
|  |  |  | Day 7 | 63 | 1 | 62 | 0 | NR | 1.32 |
|  |  |  | Day 14 | 63 | 7 | 56 | 0 | NR | 9.33 |
|  |  |  | Day 21 | 63 | 23 | 45 | 0 | 100 | 27.4 |
|  |  |  | Day 28 | 63 | 38 | 38 | 0 | 100 | 39.44 |
|  |  | pan pLDH | Day 0 | 77 | NR | NR | 0 | 100 | NR |
|  |  |  | Day 3 | 38 | 73 | 14 | 1 | 50 | 73.97 |
|  |  |  | Day 7 | NR | 74 | 11 | NR | NR | 85.53 |
|  |  |  | Day 14 | NR | 81 | 6 | NR | NR | 93.33 |
|  |  |  | Day 21 | 12 | 81 | 3 | 0 | 100 | 93.06 |
|  |  |  | Day 28 | 7 | 83 | 2 | 1 | 88.89 | 96.87 |
| Michael et al. | 2021 | HRP2 (SD Bioline) | Day 3 | 226 | 1 | 3 | 0 | 100 | 25 |
|  |  |  | Day 7 | 212 | 1 | 3 | 0 | 100 | 25 |
|  |  |  | Day 14 | 206 | 1 | 2 | 0 | 100 | 33 |
|  |  |  | Day 21 | 196 | 1 | 2 | 0 | 100 | 33 |
|  |  |  | Day 28 | 182 | 1 | 2 | 0 | 100 | 33 |
| Baiden et al. | 2012 | HRP2 (CareStart) | Day 0 | 199 | 0 | 0 | 0 | 100 | N/A |
|  |  |  | Day 1 | 174 | 78 | 2 | 4 | 97.7 | 97.5 |
|  |  |  | Day 2 | 174 | 76 | 2 | 5 | 97.7 | 97.4 |
|  |  |  | Day 3 | 174 | 73 | 3 | 5 | 97.2 | 96 |
|  |  |  | Day 7 | 152 | 67 | 9 | 7 | 95.6 | 88.2 |
|  |  |  | Day 14 | 134 | 56 | 11 | 8 | 94.4 | 83 |
|  |  |  | Day 28 | 112 | 50 | 16 | 10 | 91.1 | 75.8 |
| Biswas et al. | 2005 | HRP2 (ParaSight-F) | Day 0 | 42 | 0 | 3 | 0 | 93.3 | N/A |
|  |  |  | Day 7 | 42 | 0 | 3 | 0 | 93.3 | N/A |
|  |  |  | Day 15 | 42 | 0 | 3 | 0 | 93.3 | N/A |
|  |  |  | Day 28 | 42 | 0 | 3 | 0 | 93.3 | N/A |
